# Daily sampling of early SARS-CoV-2 infection reveals substantial heterogeneity in infectiousness

**DOI:** 10.1101/2021.07.12.21260208

**Authors:** Ruian Ke, Pamela P. Martinez, Rebecca L. Smith, Laura L. Gibson, Agha Mirza, Madison Conte, Nicholas Gallagher, Chun Huai Luo, Junko Jarrett, Abigail Conte, Tongyu Liu, Mireille Farjo, Kimberly K.O. Walden, Gloria Rendon, Christopher J. Fields, Leyi Wang, Richard Fredrickson, Darci C. Edmonson, Melinda E. Baughman, Karen K. Chiu, Hannah Choi, Kevin R. Scardina, Shannon Bradley, Stacy L. Gloss, Crystal Reinhart, Jagadeesh Yedetore, Jessica Quicksall, Alyssa N. Owens, John Broach, Bruce Barton, Peter Lazar, William J. Heetderks, Matthew L. Robinson, Heba H. Mostafa, Yukari C. Manabe, Andrew Pekosz, David D. McManus, Christopher B. Brooke

**Affiliations:** T-6, Theoretical Biology and Biophysics, Los Alamos National Laboratory, Los Alamos, NM 87545, USA; Department of Microbiology, University of Illinois at Urbana-Champaign, Urbana, IL 61801, USA; Department of Statistics, University of Illinois at Urbana-Champaign, Urbana, IL 61801, USA; Carl R. Woese Institute for Genomic Biology, University of Illinois at Urbana-Champaign, Urbana, IL 61801, USA; Department of Pathobiology, University of Illinois at Urbana-Champaign, Urbana, IL 61802, USA; Carle Illinois College of Medicine, University of Illinois at Urbana-Champaign, Urbana, IL 61801, USA; Division of Infectious Diseases and Immunology, Departments of Medicine and Pediatrics, University of Massachusetts Medical School, Worcester, MA 01655, USA; Division of Infectious Diseases, Department of Medicine, Johns Hopkins School of Medicine, Baltimore, MD 21287, USA; Division of Medical Microbiology, Department of Pathology, Johns Hopkins University School of Medicine, Baltimore, MD 21287, USA; W. Harry Feinstone Department of Molecular Microbiology and Immunology, Johns Hopkins Bloomberg School of Public Health, Baltimore, MD 21205, USA; High-Performance Biological Computing at the Roy J. Carver Biotechnology Center, University of Illinois at Urbana-Champaign, Urbana, IL 61801, USA; Veterinary Diagnostic Laboratory, University of Illinois at Urbana-Champaign, Urbana, IL 61802, USA; Center for Clinical and Translational Research, University of Massachusetts Medical School, Worcester, MA 01655, USA; UMass Memorial Medical Center, Worcester, MA 01655, USA; Department of Emergency Medicine, University of Massachusetts Medical School, Worcester, MA 01655, USA; Division of Biostatistics and Health Services Research, University of Massachusetts Medical School, Worcester, MA 01655, USA; Department of Population and Quantitative Health Sciences, University of Massachusetts Medical School, Worcester, MA 01655, USA; National Institute for Biomedical Imaging and Bioengineering, Bethesda, MD 20892, USA; Division of Cardiology, University of Massachusetts Medical School, Worcester, MA 01655, USA

## Abstract

The dynamics of SARS-CoV-2 replication and shedding in humans remain poorly understood. We captured the dynamics of infectious virus and viral RNA shedding during acute infection through daily longitudinal sampling of 60 individuals for up to 14 days. By fitting mechanistic models, we directly estimate viral reproduction and clearance rates, and overall infectiousness for each individual. Significant person-to-person variation in infectious virus shedding suggests that individual-level heterogeneity in viral dynamics contributes to superspreading. Viral genome load often peaked days earlier in saliva than in nasal swabs, indicating strong compartmentalization and suggesting that saliva may serve as a superior sampling site for early detection of infection. Viral loads and clearance kinetics of B.1.1.7 and non-B.1.1.7 viruses in nasal swabs were indistinguishable, however B.1.1.7 exhibited a significantly slower pre-peak growth rate in saliva. These results provide a high-resolution portrait of SARS-CoV-2 infection dynamics and implicate individual-level heterogeneity in infectiousness in superspreading.

## MAIN TEXT

Transmission of SARS-CoV-2 by pre-symptomatic and asymptomatic individuals has been a major contributor to the explosive spread of this virus (*1*–*5*). Recent epidemiological investigations of community outbreaks have indicated that transmission of SARS-CoV-2 is highly heterogeneous, with a small fraction of infected individuals (often referred to as “superspreaders”) contributing a disproportionate share of forward transmission (*6*–*8*). Transmission heterogeneity has also been implicated in the epidemic spread of several other important viral pathogens, including measles and smallpox (*9*). Numerous behavioral and environmental explanations have been offered to explain transmission heterogeneity, but the extent to which the underlying features of the infection process within individual hosts contribute towards the superspreading phenomenon remains unclear. Addressing this gap in knowledge will inform the design of more targeted and effective strategies for controlling community spread.

Viral infection is a highly complex process in which viral replication and shedding dynamics are shaped by the complex interplay between host and viral factors. Recent studies have suggested that the magnitude and/or duration of viral shedding in both nasal and saliva samples correlate with disease severity, highlighting the potential importance of viral dynamics in influencing infection outcomes (*10*–*13*). Variation in viral load has also been suggested to correlate with transmission risk (*14*). In addition to implications for pathogenesis and transmission, defining the contours of viral shedding dynamics is also critical for designing effective surveillance, screening, and testing strategies (*15*). To date, studies aimed at describing the longitudinal dynamics of SARS-CoV-2 shedding have been limited by (a) sparse sampling frequency, (b) failure to capture the early stages of infection when transmission is most likely, (c) absence of individual-level data on infectious virus shedding kinetics, and (d) biasing towards the most severe clinical outcomes (*16*–*21*). This is also true for viruses beyond SARS-CoV-2, as the dynamics of natural infection in humans have not been described in detail for any acute viral pathogen.

Here, we capture the early longitudinal viral dynamics of mild and asymptomatic acute SARS-CoV-2 infection in 60 people by recording daily measurements of both viral RNA shedding (from mid-turbinate nasal swabs and saliva samples) and infectious virus shedding (from mid-turbinate nasal swabs) for up to 14 days. We reveal a striking degree of individual-level heterogeneity in infectious virus shedding between individuals, thus providing a partial explanation for the central role of superspreaders in community transmission of SARS-CoV-2. We also directly compare the shedding dynamics of B.1.1.7 and non-B.1.1.7 viruses, revealing prolonged pre-peak shedding in saliva but no significant differences in nasal shedding.

Altogether, these results provide the first high-resolution, multi-parameter empirical profile of acute SARS-CoV-2 infection in humans and implicate person-to-person variation in infectious virus shedding in driving patterns of epidemiological spread of the pandemic.

## Results

### Description of cohort and study design

During the Fall of 2020 and Spring of 2021, all faculty, staff, and students at the University of Illinois at Urbana-Champaign were required to undergo at least twice weekly RTqPCR testing for SARS-CoV-2. We leveraged this large-scale, high-frequency screening program to enroll symptomatic, pre-symptomatic, and asymptomatic SARS-CoV-2-infected individuals. We enrolled university faculty, staff, and students who had a documented negative RTqPCR test result in the past seven days and were either (a) within 24 hours of a positive RTqPCR result, or (b) within five days of exposure to someone with a confirmed positive RTqPCR result. These criteria ensured that we only enrolled people within the first days of infection.

We collected both nasal and saliva samples daily for up to 14 days to generate a high-resolution portrait of viral dynamics during the early stages of SARS-CoV-2 infection. Participants also completed a daily online symptom survey. Our study cohort was primarily young (median age: 28; range: 19-73), non-Hispanic white, and skewed slightly towards males **(Table S1)**. All infections were either mild or asymptomatic, and none of the participants were ever hospitalized for COVID-19.

### Early SARS-CoV-2 infection dynamics vary significantly between individuals

To examine viral dynamics at the individual level, we plotted Ct/Cn values from both saliva and nasal samples (the RTqPCR assay used for nasal samples reports Cn rather than Ct values), Quidel SARS Sofia 2 antigen FIA results, and viral culture data from nasal swabs, as a function of time relative to observed peak in nasal viral genome load **(Fig 1A, S1)**. In many cases, we captured both the rise and fall of viral genome shedding in nasal and/or saliva samples. A comparison between individuals revealed substantial heterogeneity in shedding dynamics, with obvious differences in the duration of detectable infectious virus shedding, clearance kinetics, and the temporal relationship between shedding in nasal and saliva compartments. Further, a small subset of individuals (9 out of 60 examined) exhibited only low-level sporadic RNA shedding, with no detectable infectious virus in nasal samples.

**Figure 1:**
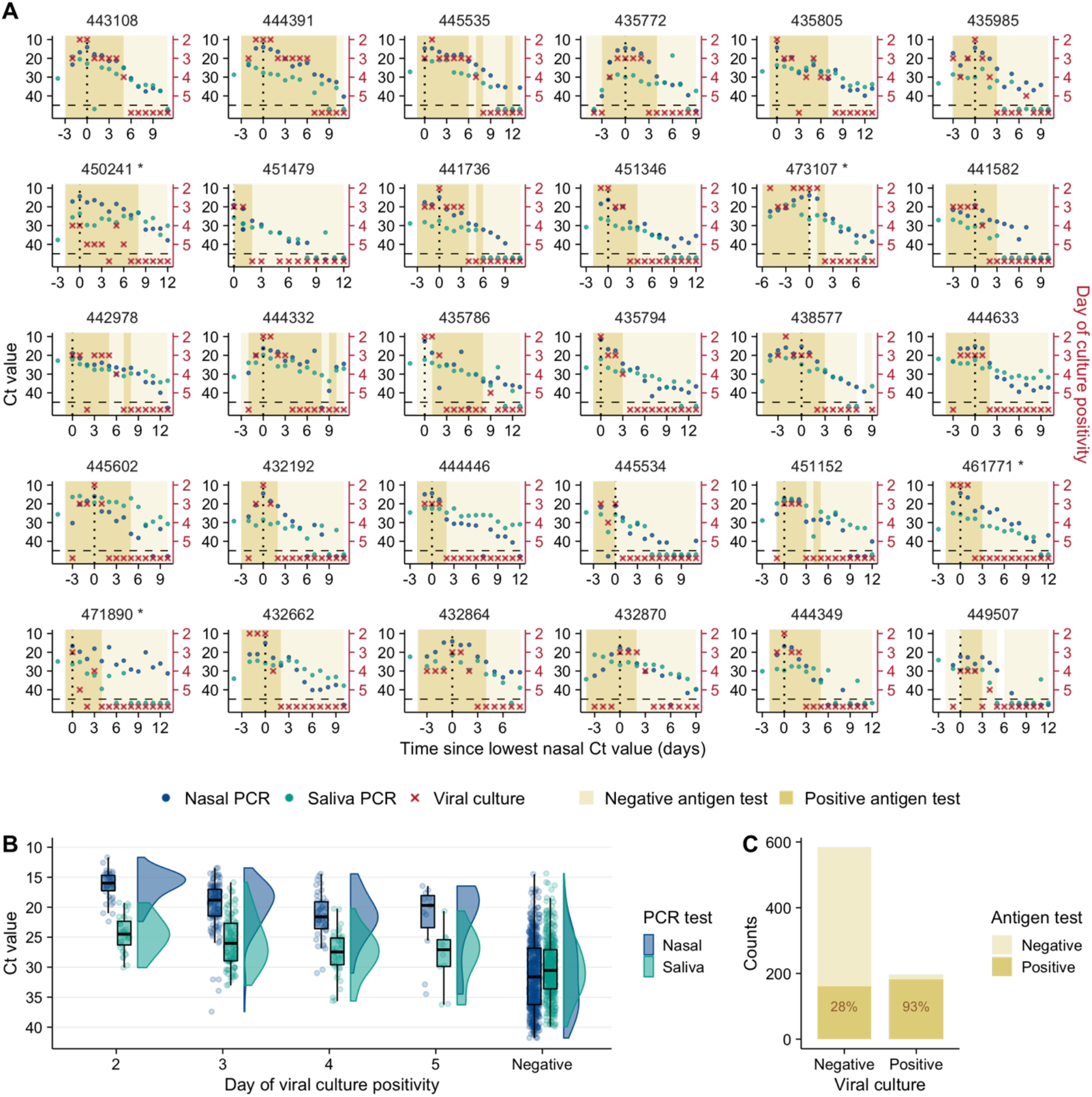
SARS-CoV-2 viral dynamics captured through daily sampling. **(A)** Temporal trends for the saliva RTqPCR (teal dots), nasal swab RTqPCR (navy blue dots), nasal swab viral culture (red crosses), and positive nasal swab antigen test results (dark mustard shaded area). The X axis shows days since the measured peak in nasal swab viral genome load, which is indicated by a vertical dotted line. The left Y axis indicates Ct values for saliva RTqPCR assay (covidSHIELD) and Cn values for nasal swab RTqPCR assay (Abbott Alinity). The right Y axis indicates results of viral culture assays, where day of culture positivity indicates the day of incubation at which > 50% of Vero-TMPRSS2 cells infected with the sample were positive for cytopathic effect. Horizontal dashed line indicates limit of detection of RTqPCR and viral culture assays. Title of each plot indicates the participant ID for the top 30 individuals with the most data points (the remaining 30 participants are shown in figure S1). Single asterisk next to the participant ID indicates B.1.1.7 variant infection. **(B)** Individual Ct (for saliva) and Cn (for nasal swabs) values from samples plotted based on concurrent results from the viral culture assay. Negative indicates samples for which viral culture assay showed no viral growth after 5 days. Note the Y axes are reversed in panel (A) and (B). **(C)** Plot showing antigen FIA results from days where participants tested either positive or negative by viral culture. The text inside the bars indicates the percentage of antigen FIA results that were positive when concurrent viral culture sample was positive or negative.

Generally, earlier positivity results in the viral culture assay (which suggests higher infectious viral loads) were associated with lower Ct values in nasal samples **(Fig 1B)**. This is unsurprising, as both nasal viral genome load and viral infectivity were assayed using the same sample. Saliva Ct values tended to be higher than matched nasal samples, likely due in part to the lower molecular sensitivity of the specific saliva RTqPCR assay used which does not include an RNA extraction step (*22*). For both sample types, the relationship between viral culture results and Ct values was not absolute, as several nasal swab samples with Ct values greater than 30 also tested positive for infectious virus. These data indicate that caution must be exercised when using a simple Ct value cutoff as a surrogate for infectious status.

We also assessed the relationship between antigen FIA and viral culture results and found that participants tested positive by antigen FIA on 93% of the days on which they also tested positive by viral culture **(Fig 1C)**. This finding is consistent with earlier cross-sectional studies examining the relationship between antigen tests positivity and infectious virus shedding (*23, 24*).

While the symptom profiles self-reported by study participants varied widely across individuals, all cases were mild and did not require medical treatment **(Fig S2)**. To determine whether any specific symptoms correlated with viral culture positivity, we compared the reported frequencies for each symptom on days where individuals tested viral culture positive or negative **(Fig S3)**. Muscle aches, runny nose, and scratchy throat were significantly more likely to be reported on days when participants were viral culture positive, suggesting these specific symptoms as potential indicators of infectious status. No other symptoms examined exhibited a clear association with viral culture status. Self-reported symptom data from this study may be partially skewed by having been collected after participants were notified of their initial positive test result or potential exposure.

### Within-host mechanistic models capture viral dynamics in nasal and saliva samples

To better quantify the specific features of viral dynamics within individuals, we implemented five within-host models based on models developed previously for SARS-CoV-2 and influenza infection (see Supporting Text for details) **(Figs 2A, S4)** (*25, 26*). We fit these models to viral genome loads derived from RTqPCR Ct/Cn values using a population mixed effect modeling approach (see Supporting Text). The viral dynamics in nasal and saliva samples were distinct from each other in most individuals, indicating strong compartmentalization of the oral and nasal cavities. We thus fit the models to data from nasal and saliva samples separately. For either nasal or saliva samples, we selected data from 56 out of 60 individuals for model fitting. For each sample type, data from 4 individuals were excluded because viral genome loads were very low throughout the sampling period.

**Figure 2:**
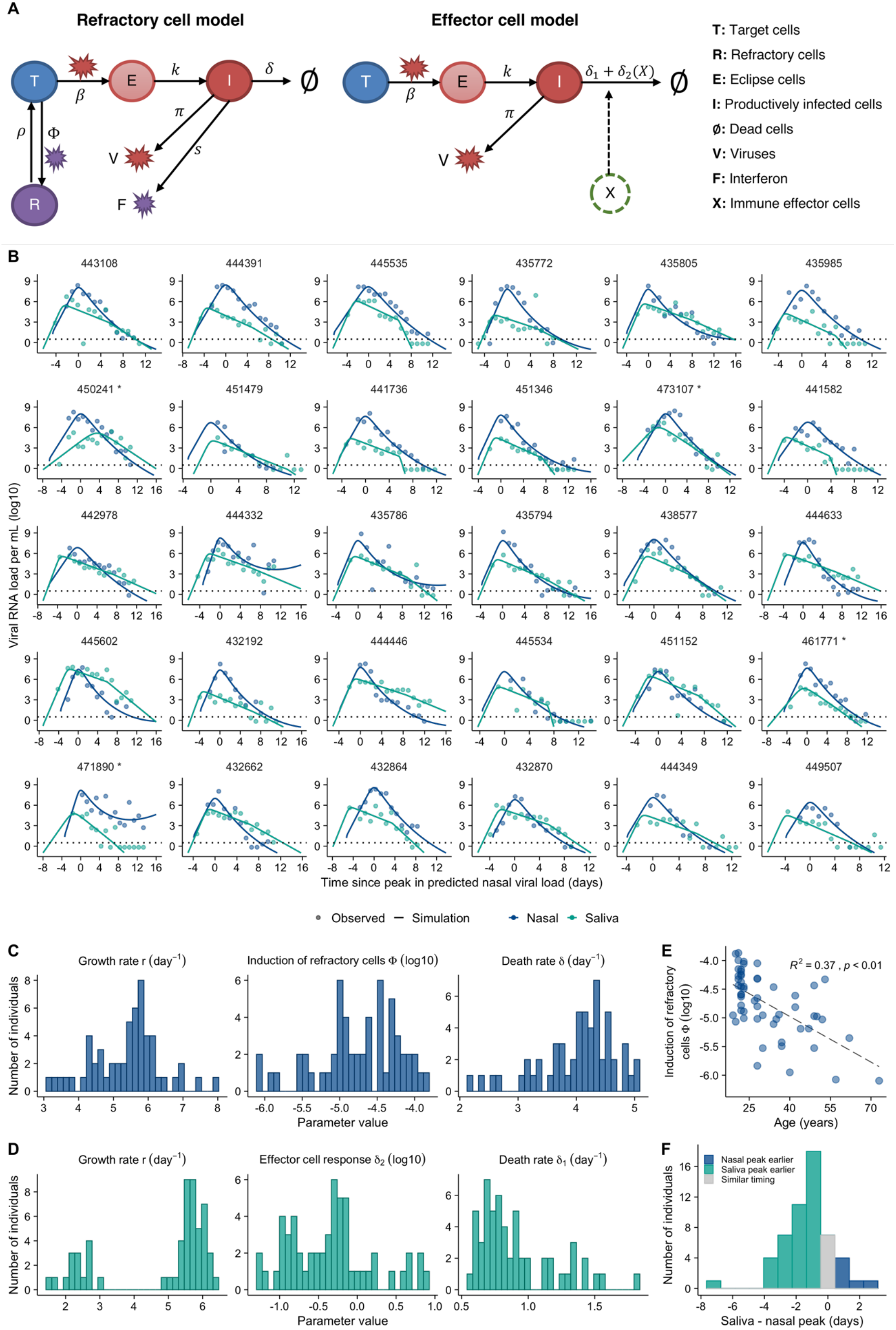
Model fits quantify heterogeneity in viral dynamics and discordance in genome shedding between nasal and saliva samples. **(A)** Diagrams outlining structures of the refractory cell model and the immune effector cell model that best fit the nasal swab and saliva RTqPCR data, respectively. In the refractory cell model, target cells (T) are infected by viruses (V) at rate β. Infected cells first become eclipse cells (E) and do not produce viruses; at rate κ, eclipse cells become productively infected cells (I) producing both viruses and interferon (F) at rate πand s, respectively. They die at rate δ. Binding of interferons with target cells induces an antiviral response that converts target cells into cells refractory to infection (R). The rate of induction of the antiviral response is Ф. Refractory cells can revert to target cells at rate р. In the effector cell model, instead of modeling the interferon response explicitly, we assume that over the course of infection, immune effector cells (X) that clear infected cells are activated and recruited. This leads to an increase in infected cell death rate from δ_1_ to δ_1_ + δ_2_ **(B)** Model fits to nasal sample (navy blue) and saliva (teal) RTqPCR results from the same subset of individuals shown in figure 1A. Dotted lines represent the limit of detection for the RTqPCR assays. The Y axis is shown on a log 10 scale. **(C)** Distributions of key individual parameter estimates from the refractory cell model (nasal data) across individuals in this cohort. **(D)** Distributions of key individual parameter estimates from the immune effector cell model (saliva data). **(E)** Association between age and the estimated strength of the antiviral immune response (Φ) based on nasal sample data. The Y axis is shown on a log 10 scale. **(F)** Distribution of the differences in the estimated times of peak viral genome loads between the saliva and nasal samples. Nasal sample peak is set to time=0 for all participants. Bars colored teal and navy blue represent estimated saliva peaks that occur at least 0.5 day earlier and later than nasal samples, respectively. The gray bar indicates the number of individuals that have similar timing in the peaks.

To identify factors that might partially explain the observed variation in individual-level dynamics, for each model we tested whether the age of the participants or the infecting viral genotype (*i.e*. non-B.1.1.7 vs. B.1.1.7) co-vary with any of the estimated model parameters in the model fitting. A total of 107 model variations were tested (see Supporting text for the testing strategy). We compared the relative abilities of these model variations to capture the RTqPCR data using the Akaike Information Criterion (AIC) and found that in general, the refractory model and the effector cell model best describe data from nasal and saliva samples, respectively **(Table S2, S3)**. In the refractory model **(Fig 2A)**, we assumed that target cells can be rendered refractory to infection through the activity of soluble immune mediators released by infected cells, such as interferon (*27*). In the best-fit immune effector cell model **(Fig 2A)**, we assumed that innate and adaptive immune cells are activated and recruited to eliminate infected cells, leading to increased viral clearance (*26*). We estimated that all fitted parameters with a random effect differ between non-B.1.1.7 and B.1.1.7 infections. See **Tables S4, S5 and S6** for estimated values of the population and individual parameters and the fixed parameter values, respectively. Overall, these models described the viral genome load data in both nasal and saliva samples very well **(Fig 2B)**.

The frequent longitudinal sampling of participants during early infection provided a unique opportunity to precisely quantify viral load kinetics during the viral expansion phase, prior to the peak in genome shedding. We estimated the mean exponential expansion rate *r* (growth rate, for short) to be 5.4/day (SD: ±0.98/day) in nasal swabs and 4.8/day (SD: ±1.6/day) in saliva **(Fig 2C-D)**. Note that as we will show below, the growth rate of the B.1.1.7 variant is lower than that of non-B.1.1.7 viruses in saliva, leading to a bimodal distribution **(Fig 2D)**.

Viral clearance kinetics clearly differed between nasal and saliva samples **(Fig 2B-D)**. For nasal samples, viral genome loads decreased rapidly after peak, with an estimated death rate of productively infected cells at 4.0/day (SD: ±0.6/day); however, the rate of viral decline decreased over time. In saliva, post-peak viral genome loads declined initially at a slower rate on average (0.9/day; SD: ±0.3/day) than the decline in nasal samples. However, our model suggested the existence of a second clearance phase with a more rapid decline, potentially due to the onset of effector cell and/or neutralizing antibody responses.

Interestingly, the model predicts a significant correlation (p<0.01) in nasal samples between age and the Φ parameter which describes the effectiveness of the anti-viral immune response in rendering target cells refractory to infection (**Fig 2E)**. This suggests that innate immune responses are less effective at limiting SARS-CoV-2 in the nasal compartment of older individuals within our cohort, consistent with prior studies describing dysregulation of innate immunity to viral infection in aged individuals (*28*–*30*). There was no significant correlation between age and either growth rate or clearance rate in nasal samples. In saliva, none of the individual-level model parameter estimates correlated with participant age **(Fig S5)**.

Overall, we noted a surprising degree of discordance in viral dynamics between nasal and saliva samples for many participants. In most individuals (41 out of 54 analyzed), viral genome shedding peaked at least 1 day earlier in saliva than in nasal samples **(Fig 2F)**. In contrast, the peak in nasal shedding only preceded the saliva peak by at least 1 day in 3 individuals.

### Significant heterogeneity in the infectious potential of individuals

We next examined the duration of infectious virus shedding in nasal samples, as a surrogate for the infectious potential of an individual; 51 out of 60 individuals tested positive by viral culture on nasal swabs at least once during the sampling period **(Fig 3A)**. We found a weak positive correlation between the duration of viral culture positivity and participant age **(Fig 3B)**. Of note, many study participants were viral culture positive on the first day of sample collection, suggesting that we failed to capture the onset of viral culture positivity for these individuals and thus may be underestimating the duration of infectious virus shedding for a subset of study participants.

**Figure 3:**
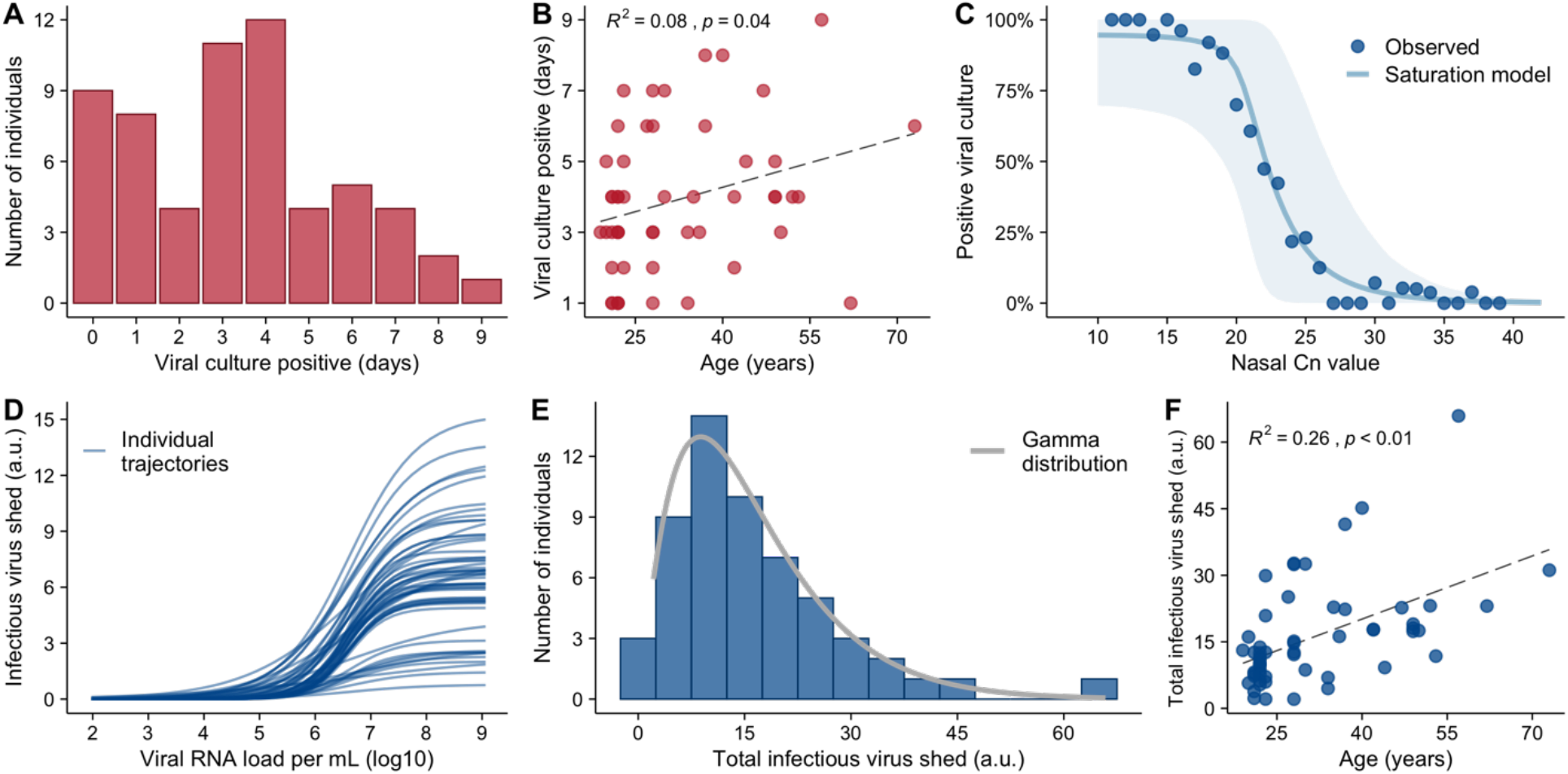
Substantial heterogeneity in infectious virus shedding between individuals. **(A)** The distribution in the numbers of days in which participants tested positive by viral culture on nasal swabs following study enrollment. **(B)** Association between age of study participants and the number of days of positive viral culture. **(C)** Plot shows the relationship between the Cn value in nasal samples and the probability of the sample being viral culture positive summarized across all individuals. Individual samples were binned based on their Cn values. Dots indicate the observed percentage of positive samples within a bin that were viral culture positive. The solid line and the shaded area are the mean and the 90% confidence intervals, respectively, of the trajectories generated using the best-fit parameters of the saturation model (see Fig S7 for individual fits). **(D)** The relationship between infectious virus shed (in arbitrary units) and viral genome load predicted by the saturation model for 56 individuals in our analysis. **(E)** Distribution of the estimated total cumulative level of infectious virus (in arbitrary units) shed from the nasal passage by each participant over the course of infection. Solid line shows the best-fit gamma distribution with a shape parameter of 2. **(F)** Association between age and the estimated total infectious virus (in arbitrary units) shed by each individual. The R^2^ value and the p value from a linear regression (dashed line) are shown in panels (B) and (F).

To better quantify the infectious potential of each individual, we first used the viral culture data as a measure for intrinsic infectiousness (infectiousness for short below) to characterize how infectiousness depends on the viral genome load. We fitted three alternative models as proposed before (*31*) to the paired nasal RTqPCR and viral culture data collected from each individual using a non-linear mixed effect modeling approach (see **Fig. S6** for a workflow and Supporting Text for more details). Comparing models using the AIC scores, we found that the relationship is best described by a saturation model where the infectious virus load is a Hill-type function of the viral genome load **(Fig 3C, S7 and Table S7)**. See **Table S8** for the best-fit parameter values.

Using the best-fit model, we estimated the infectiousness of each individual over the course of infection (**Fig. S8**). Note that the dataset only allows us to estimate a quantity that is a constant proportion of the infectious virus load (rather than its absolute value) across time and between individuals, thus we report the predicted the values in arbitrary units (a.u.) as a relative measure of infectiousness. Our model predicts that infectious virus shedding increases sharply when viral genome loads rise above approximately 10^5^ to 10^6^ copies/mL (**Fig. 3D**). Importantly, there exists a high level of heterogeneity in infectiousness across different individuals that is not fully explained by differences in viral genome load (**Fig. 3D**). For example, at a viral genome load of 10^8^ copies/mL, infectious virus shedding is higher than 10 a.u. in some individuals; while in other individuals it is below 1 a.u. This suggests that viral genome loads are not precisely predictive of infectiousness.

We next estimated the total infectiousness of each individual by integrating the area under the infectious virus load curve over the course of infection. This approach again revealed a large degree of heterogeneity in individual-level infectiousness, with a more than 30-fold difference between the highest and lowest estimated infectiousness (65.9 and 2.1 a.u., respectively) **(Fig 3E)**. These data suggest that the previously reported heterogeneity in secondary transmission rates (*6, 7*) can arise from heterogeneity in intrinsic infectiousness in addition to heterogeneity in contact structure. We then fit a gamma distribution to the individual-level infectiousness data and found that a distribution with a shape parameter of 2 describes the majority of the individuals well, with the exception of one individual who had the highest level of infectiousness **(Fig 3E)**. This again emphasizes the potential for a small subset of individuals that exhibit high intrinsic infectiousness to function as ‘superspreaders’ if they have frequent and/or high-risk contacts during the infectious period. Finally, we observed a significant correlation between age and total infectiousness (p<0.01; R^2^ = 0.26, **Fig 3F)**.

### Analysis of B.1.1.7 (Alpha) viral dynamics

Finally, we asked whether infection with the B.1.1.7 (Alpha) variant of concern (VOC) was associated with any significant differences in viral dynamics that could potentially explain the enhanced transmissibility of this genotype (*32*–*34*). Previous studies have suggested that B.1.1.7 infection may result in higher peak viral loads or prolonged shedding compared with other circulating genotypes (*35*–*37*). Within our cohort, 15 out of 60 individuals were infected with B.1.1.7.

Both the empirical data and our model analysis suggest that viral genome shedding dynamics in nasal samples are indistinguishable between B.1.1.7 and non-B.1.1.7 infections (none of the latter were VOC genotypes with the exception of a single P.1 (Gamma) infection; see **Table S9**), and that there is no significant difference between B.1.1.7 and non-B.1.1.7 viruses in total infectiousness predicted by our model **(Fig 4A-B)**. Comparison of genome shedding in saliva between B.1.1.7 and non-B.1.1.7 infected individuals revealed a slower rise in pre-peak viral loads for B.1.1.7 versus non-B.1.1.7 **(Fig 4C)**. This is corroborated by the mathematical modeling results, where viral growth rates were estimated to be significantly slower for B.1.1.7 compared with non-B.1.1.7 viruses (p-value <0.01) **(Fig 4D)**. No clear differences in peak viral genome loads or overall clearance kinetics in saliva were apparent, although the peak viral genome loads for B.1.1.7 seemed to be slightly, but not significantly higher (p=0.2). With a slower rise to peak viral load for B.1.1.7 and similar peak viral loads between B.1.1.7 and non-B.1.1.7 viruses, our model predicts a longer pre-peak shedding period for B.1.1.7. These data implicate either the delayed rise in pre-peak shedding in saliva or other mechanisms not reflected in viral dynamics in the increased transmissibility of this variant.

**Figure 4:**
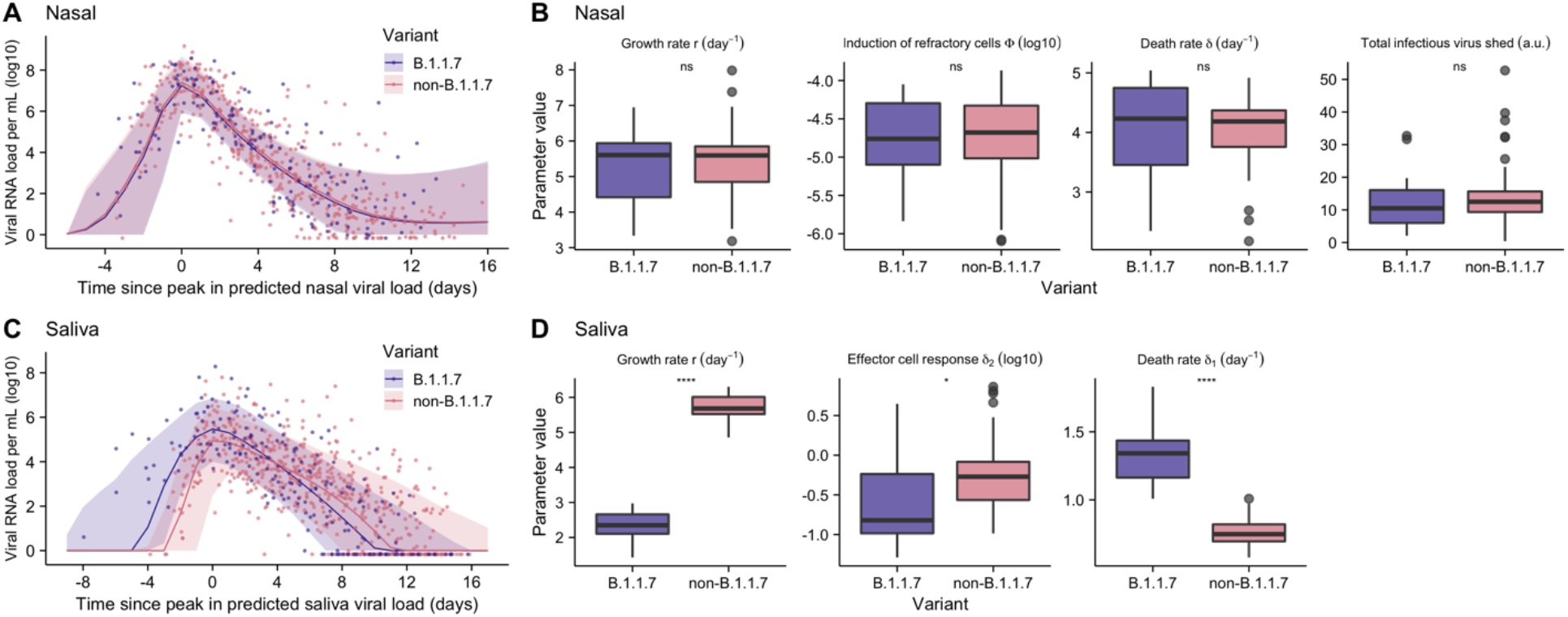
Comparison of viral dynamics between B.1.1.7 and non-B.1.1.7 viruses. **(A, C)** Viral genome load of B.1.1.7 infections (purple) and non-B.1.1.7 infections (pink) over time in (A) nasal and (C) saliva samples, as measured by RTqPCR. Dots indicate individual viral genome load measurements. Ribbons indicate 90% confidence intervals of viral genome load trajectories generated using population parameters estimated from modeling analysis (Table S4). **(B, D)** Estimated values for the indicated model parameters from individual B.1.1.7 and non-B.1.1.7 infections (see Table S5 for parameter values). Total infectious virus shed after adjusting for age is shown in panel (B) (see Methods).

## Discussion

This study describes the results of daily multi-compartment sampling of viral dynamics within dozens of individuals newly infected with SARS-CoV-2, and provides the most comprehensive, high-resolution description of SARS-CoV-2 (or any acute RNA virus) shedding and clearance dynamics in humans to date.

Superspreading, in which a small subset of infected individuals are responsible for a disproportionately large share of transmission events, has been identified as a major driver of community spread of SARS-CoV-2, SARS-CoV, and many other acute viral pathogens (*6, 7, 9*). Superspreading is believed to arise from heterogeneity in both (1) contact structure between individuals arising from behavioral and environmental factors, and (2) the intrinsic infectiousness of individuals (*9, 38*). While heterogeneity in contact structure has been studied extensively (*39*– *42*), the extent of heterogeneity in infectiousness arising from individual-level infection processes has not been empirically measured during natural human infection. As a result, the extent to which individual-level heterogeneity in viral dynamics contributes to superspreading remains unknown.

To address this question, we empirically quantified infectious virus shedding through daily longitudinal sampling of individuals infected with SARS-CoV-2. The substantial heterogeneity in infectious virus shedding that we observed among individuals indicates that superspreading is likely driven by individual-level variation in specific features of the infection process, in addition to behavioral and environmental factors. We also found that heterogeneity in infectious virus shedding was only partly explained by individual-level heterogeneity in viral genome load dynamics, suggesting that additional factors such as variation in the timing and magnitude of the neutralizing antibody response might contribute (*43*). Our results here suggest caution in assessing the infectiousness of an individual using viral genome load data alone. Further, the absence of clear viral genetic correlates of infectiousness within this dataset suggests the existence of specific host determinants of superspreading potential. While we identified age as a significant correlate of infectiousness, additional determinants likely exist. Identification of these correlates could aid future efforts to mitigate community spread of the virus.

Our finding that viral shedding often peaks earlier in saliva versus the nasal compartment, sometimes by several days, corroborates a recent study of four individuals (*44*) and has several important implications. First, saliva screening may be a more effective sample type than nasal swabs for detecting infected individuals prior to or early in the infectious period (*45*). Early detection and isolation of infected individuals is absolutely critical for breaking transmission chains (*15*). Moreover, early viral shedding from the oral cavity may contribute to the high prevalence of pre-symptomatic SARS-CoV-2 transmission. We were unable to directly assess viral infectivity in saliva so it remains unclear whether the earlier peaks in viral RNA shedding that we observed in saliva reflect earlier shedding of transmission-competent virus. The earlier detection of virus in saliva also raises questions about the initial site of SARS-CoV-2 infection. A recent study demonstrated that both salivary glands and oral mucosal epithelium can support SARS-CoV-2 replication, suggesting that infection could be initiated within the oral cavity (*46*). Alternatively, if infection is initiated in the nasopharynx or soft palette, viral RNA might be detectable in saliva prior to detection in the mid-turbinate swabs used in this study. The discordance in shedding dynamics between oral and nasal samples that we observed in many participants is consistent with a significant degree of compartmentalization between these adjacent but distinct tissue sites, as has been observed in animal models of influenza virus infection (*47*).

The specific mechanisms driving the enhanced transmissibility of the B.1.1.7 variant remain poorly understood. Recent studies have identified alterations in the structural conformation of the spike protein and enhanced antagonism of innate immunity by B.1.1.7 as potential contributors (*48, 49*). Contrary to previous clinical studies, we observed no significant differences in either peak viral loads or clearance kinetics between B.1.1.7 and non-B.1.1.7 viruses as measured in either nasal swabs or saliva. Previous reports of differences in Ct values or duration of shedding were based on either small sample sizes (*36*) or cross-sectional analyses of individuals who sought testing and were thus likely symptomatic at the time of sampling (*21*). Our results are consistent with studies demonstrating the absence of a growth advantage for B.1.1.7 in primary human respiratory epithelial cells (*50, 51*). If the timing of symptom onset differs between B.1.1.7 and non-B.1.1.7 infections, it could potentially explain why cross-sectional analyses of viral loads might register lower Ct values for B.1.1.7 samples. Interestingly, we did observe a significantly slower pre-peak growth rate of B.1.1.7 than non-viruses in saliva but not nasal swabs, resulting in more prolonged shedding in saliva prior to peak viral load. Because symptom onset seems to occur around the time of peak viral load (*1, 25, 52*) the prolonged pre-peak shedding in saliva by B.1.1.7 could potentially enhance transmissibility by extending the pre-symptomatic shedding period of the virus, however more data are needed to determine whether there is a mechanistic link between pre-peak shedding kinetics and transmission potential. Alternatively, the enhanced transmissibility of the B.1.1.7 variant may also be driven by features not reflected in shedding dynamics, *e.g*. enhanced environmental stability or a lower infectious dose threshold.

Altogether, our data provide an unprecedented view of the longitudinal viral dynamics of SARS-CoV-2 infection in humans and implicate both individual-level heterogeneity in viral shedding and the oral compartment and as playing critical roles in transmission.

## Methods

This study was approved by the Western Institutional Review Board, and all participants provided informed consent.

### Participants

All on-campus students and employees of the University of Illinois at Urbana-Champaign are required to submit saliva for RTqPCR testing every 2-4 days as part of the SHIELD campus surveillance testing program. Individuals testing positive were instructed to isolate and were eligible to enroll in this study for a period of 24 hours following receipt of their positive test result. Close contacts of individuals who test positive (particularly those co-housed with them) are instructed to quarantine and were eligible to enroll for up to 5 days after their last known exposure to an infected individual. All participants were also required to have received a negative saliva RTqPCR result 7 days prior to enrollment.

Individuals were recruited via either a link shared in an automated text message providing isolation information sent within 30 minutes of a positive test result, a call from a study recruiter, or a link shared by an enrolled study participant or included in information provided to all quarantining close contacts. In addition, signs were used at each testing location and a website was available to inform the community about the study.

Participants were required to be at least 18 years of age, have a valid university ID, speak English, have internet access, and live within 8 miles of the university campus. After enrollment and consent, participants completed an initial survey to collect information on demographics and health history and were provided with sample collection supplies. Participants who tested positive prior to enrollment or during quarantine were followed for up to 14 days. Quarantining participants who continued to test negative by saliva RTqPCR were followed for up to 7 days after their last exposure. All participants’ data and survey responses were collected in the Eureka digital study platform.

### Sample collection

Each day, participants were remotely observed by trained study staff collecting:

1. 2 mL of saliva into a 50mL conical tube.
2. 1 nasal swab from a single nostril using a foam-tipped swab that was placed within a dry collection tube.
3. 1 nasal swab from the other nostril using a flocked swab that was subsequently placed in a collection vial containing viral transport media (VTM).

The order of nostrils (left vs. right) used for the two different swabs was randomized. For nasal swabs, participants were instructed to insert the soft tip of the swab at least 1 cm into the indicated nostril until they encountered mild resistance, rotate the swab around the nostril 5 times, leaving it in place for 10-15 seconds. After daily sample collection, participants completed a symptom survey. A courier collected all participant samples within 1 hour of collection using a no-contact pickup protocol designed to minimize courier exposure to infected participants.

### Saliva RTqPCR

After collection, saliva samples were stored at room temperature and RTqPCR was run within 12 hours of initial collection. The protocol for the covidSHIELD direct saliva-to-RTqPCR assay used has been detailed previously (*22*). In brief, saliva samples were heated at 95°C for 30 minutes, followed by the addition of 2X Tris/Borate/EDTA buffer (TBE) at a 1:1 ratio (final concentration 1X TBE) and Tween-20 to a final concentration of 0.5%. Samples were assayed using the Thermo Taqpath COVID-19 assay.

### Antigen testing

Foam-tipped nasal swabs were placed in collection tubes, transported with cold packs, and stored at 4°C overnight based on guidance from the manufacturer. The morning after collection, swabs were run through the Sofia SARS antigen FIA on Sofia 2 devices according to the manufacturer’s protocol.

### Nasal swab RTqPCR

Collection tubes containing VTM and flocked nasal swabs were stored at -80°C after collection and were subsequently shipped to Johns Hopkins University for RTqPCR and virus culture testing. After thawing, VTM was aliquoted for RTqPCR and infectivity assays. One ml of VTM from the nasal swab was assayed on the Abbott Alinity per manufacturer’s instructions in a College of American Pathologist and CLIA-certified laboratory. Calibration curve for Alinity assay was determined using digital droplet PCR (ddPCR) as previously described (*53*).

### Virus culture from nasal swabs

Vero-TMPRSS2 cells were grown in complete medium (CM) consisting of DMEM with 10% fetal bovine serum (Gibco), 1 mM glutamine (Invitrogen), 1 mM sodium pyruvate (Invitrogen), 100 U/ml of penicillin (Invitrogen), and 100 μg/ml of streptomycin (Invitrogen) (*54*). Viral infectivity was assessed on Vero-TMPRSS2 cells as previously described using infection media (IM; identical to CM except the FBS is reduced to 2.5%) (*24*). When a cytopathic effect was visible in >50% of cells in a given well, the supernatant was harvested. The presence of SARS-CoV-2 was confirmed through RTqPCR as described previously by extracting RNA from the cell culture supernatant using the Qiagen viral RNA isolation kit and performing RTqPCR using the N1 and N2 SARS-CoV-2-specific primers and probes in addition to primers and probes for human RNaseP gene using synthetic RNA target sequences to establish a standard curve (*55*).

### Viral genome sequencing and analysis

Viral RNA was extracted from 140 uL of heat inactivated (30 minutes at 95°C, as part of protocol detailed in (*22*)) saliva samples using the QIAamp viral RNA mini kit (QIAGEN). 100ng of viral RNA was used to generate cDNA using the SuperScript IV first strand synthesis kit (Invitrogen). Viral cDNA was then used to generate sequencing libraries using the Swift SNAP Amplicon SARS CoV2 kit with additional coverage panel and unique dual indexing (Swift Biosciences) which were sequenced on an Illumina Novaseq SP lane. Data were run through the nf-core/viralrecon workflow (https://nf-co.re/viralrecon/1.1.0), using the Wuhan-Hu-1 reference genome (NCBI accession NC_045512.2). Swift v2 primer sequences were trimmed prior to variant analysis from iVar version 1.3.1 (https://genomebiology.biomedcentral.com/articles/10.1186/s13059-018-1618-7) retaining all calls with a minimum allele frequency of 0.01 and higher. Viral lineages were called using the Pangolin tool (https://github.com/cov-lineages/pangolin) version 2.4.2, pango version 1.2.6, and the 5/19/21 version of the pangoLEARN model; based on the nomenclature system described in (*56*).

### Mathematical modeling and statistical analyses

See **Fig S6** for an overview of the modeling and analysis workflow of various datasets. See supplementary material for descriptions of the viral dynamic models, models describing the relationship between the infectious viral load and the viral genome load, model fitting and model comparison procedures.

Associations between the age of individuals and each parameter of interest (on a linear or a log scale) are examined using a linear regression analysis. The R-squared value and the p-value are reported. The difference in the distribution of a parameter of interest between the non-B.1.1.7 infection group and the B.1.1.7 infection group is assessed using a univariate analysis and the p-value is calculated using the Mann-Whitney test. The comparison of infectious virus shedding between the non-B.1.1.7 infection group and the B.1.1.7 infection group is performed using a multivariate analysis with age as an additional variate. The predicted levels of infectious viral shedding after adjusting for age by assuming age is at 28, i.e. the median age of the cohort, are shown in Fig. 4C.

## Data Availability

Due to institutional (LANL) policies, the codes for the viral dynamic models cannot be made publicly available. The simulation results of the models can be fully reproduced from the equations in the Supporting Text and the parameter values listed in Supplementary Tables. Raw data will be made available at the time of publication.

## Acknowledgments

We wish to thank Shumon Ahmed, Carly Bell, Nate Bouton, Callie Brennen, Justin Brown, Coleco Buie, Emmaline Cler, Gary Cole, Trey Coleman, Alastair Dunnett, Lauren Engels, Savannah Feher, Kelsey Fox, Lexi Freeman, Yesenia Gonzalez, Montez Harris, Darcy Henness, Dan Hiser, Ayeshah Hussain, Daryl Jackson, Junko Jarrett, Michael Jenkins, Kalombo Kalonji, Syntyche Kanku, Steven Krauklis, Mary Krouse, Elmore Leshoure, Joe Lewis, Maggie Li, Angel Lopez, Guadalupe Lopez, Emily Luna, Chun Huai Luo, Colby Mackey, Skyler McLain, Yared Berhanu Melesse, Madison O’Donnell, Savanna Pflugmacher, Denver Piatt, Skyler Pierce, Gina Quitanilla, Ameera Samad, MacKenzie Scroggins, Monique Settles, Macie Sinn, Pete Varney, Evette Vlach, Raeshun Williams-Chatman, and Todd Young for their efforts supporting recruitment, enrollment, logistics, and/or sample collection and processing. We also thank Jeffrey Olgin, Noah Peyser, and Xochitl Butcher for assistance with the Eureka platform, Michelle Lore for assistance with REDcap, Melanie Loots for assistance with administration, Gillian Snyder for assistance in development of study protocols and logistics, and Erin Iturriaga and Jue Chen for study protocol development. Finally, we are grateful to Dr. Alvaro Hernandez and Ms. Chris Wright of the DNA Services Lab within the Roy J. Carver Biotechnology Center for expert assistance in establishment of a SARS-CoV-2 genomic sequencing protocol. Vero-TMPRSS2 cells were kindly provided by the National Institute of Infectious Diseases, Japan. Sofia 2 devices and associated supplies were provided to Carle Foundation Hospital by Quidel, however Quidel played no role in the design of the study or the interpretation or presentation of the data.

## Author contributions

Conceptualization: RK, RLS, WJH, YCM, AP, LLG, CBB

Data curation: RLS, BB, PL

Formal Analysis: RK, PPM, RLS

Funding acquisition: DDM, LLG

Investigation: RK, PPM, RLS, AM, MC, NG, CHL, JJ, AC, TL, MF, KW, CJF, LW, RF, MEB, KKC, HC, KRS, ANO, JB, MLR

Methodology: RK, RLS, CBB

Project administration: DCE, KRS, SB, SLG, CR, JY, JQ

Software: RK, PPM, RLS

Supervision: HHM, YCM, AP, LLG, CBB

Visualization: RK, PPM, RLS, CBB

Writing – original draft: RK, CBB

Writing – review & editing: RK, PPM, RLS, YCM, AP, LLG, CBB

## Supplemental figure legends

**Fig S1: Remainder of individual plots**. Plots of longitudinal assay results from study participants not shown in Figure 1A. Single asterisk next to the participant ID indicates B.1.1.7 variant infection, while double asterisks indicate P1 variant infection.

**Fig S2: Individual-level symptom data**. Self-reported symptom data from study participants, overlaid with viral culture status. Participants were asked to complete a survey through the Eureka digital study platform inquiring about the presence or absence of the indicated set of symptoms each day after sample collection.

**Fig S3: Comparison of symptoms and viral culture status**. Plots show the proportions of either viral culture negative or viral culture positive days for which participants reported the indicated symptoms. Data are only shown for individuals who reported the indicated symptom at least once.

**Fig S4: Model structures**. Diagrams showing the structures of the additional three models (not shown in Figure 2A) considered for describing viral load data. See Supporting Text for descriptions of the models.

**Fig S5: Model parameter estimates as a function of age**. Plots showing the relationship between age and the indicated model parameter estimates for **(A)** the refractory cell model (nasal data) and the **(B)** the immune effector cell model (saliva data). Linear regressions were performed on the data. R^2^ values and p-values are shown.

**Fig S6: Analysis workflow**. Diagram indicating how empirical RTqPCR and viral culture data were used to generate estimations of individual level viral dynamics and infectiousness.

**Fig S7: The saturation model accurately predicts the cell culture positivity data**. Lines denote the predicted probability of cell culture being positive. Dots denotes cell culture positivity data, where a dot is at 1 or 0 when the cell culture is positive or negative, respectively.

**Fig S8: Individual infectiousness plots**. Estimated infectiousness over time plotted for individual study participants. Dashed lines indicate inferred peak in infectiousness.

## Supplemental tables

**Table S1: Study cohort demographics data**

**Table S2: Comparing model fits to longitudinal viral load data from nasal swab samples using AIC scores**

**Table S3: Comparing model fits to longitudinal viral load data from saliva samples using AIC scores**

**Table S4: Estimated population parameter values from the best models**

**Table S5: Individual parameter values estimated from the best fit models**

**Table S6: The fixed parameters and their values in all viral dynamic models**

**Table S7. Model comparison using AIC scores of three models describing the relationship between the viral genome load and cell culture positivity**

**Table S8. The estimated parameters in the best-fit model, i.e. the saturation model, to the paired RTqPCR and cell culture positivity data**

**Table S9: Viral genotype data**

