## Supplemental Figures for "Daily sampling of early SARS-CoV-2 infection reveals substantial heterogeneity in infectiousness"

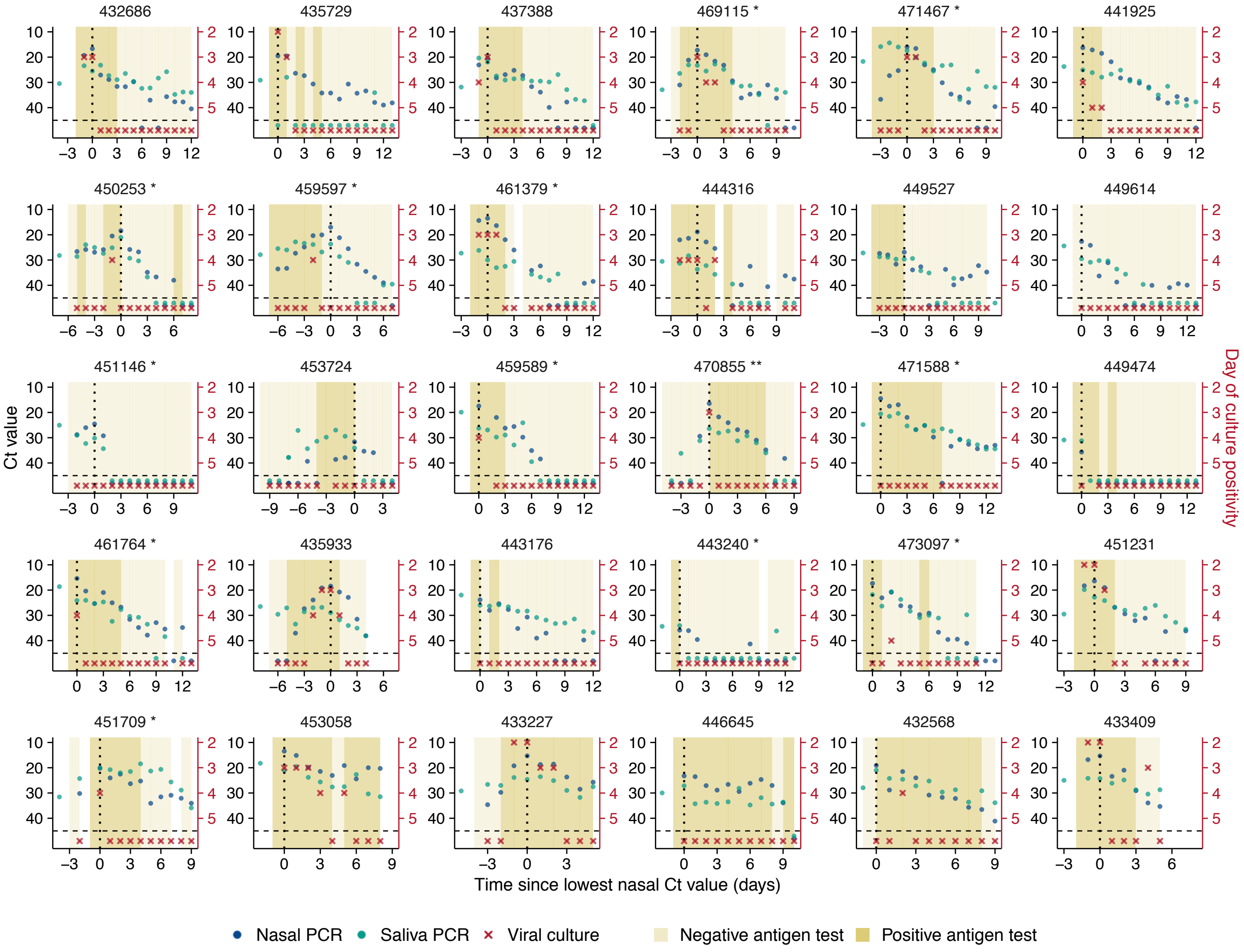


**Fig S1: Remainder of individual plots.** Plots of longitudinal assay results from study participants not shown in Figure 1A. Single asterisk next to the participant ID indicates B.1.1.7 variant infection, while double asterisks indicate P1 variant infection.

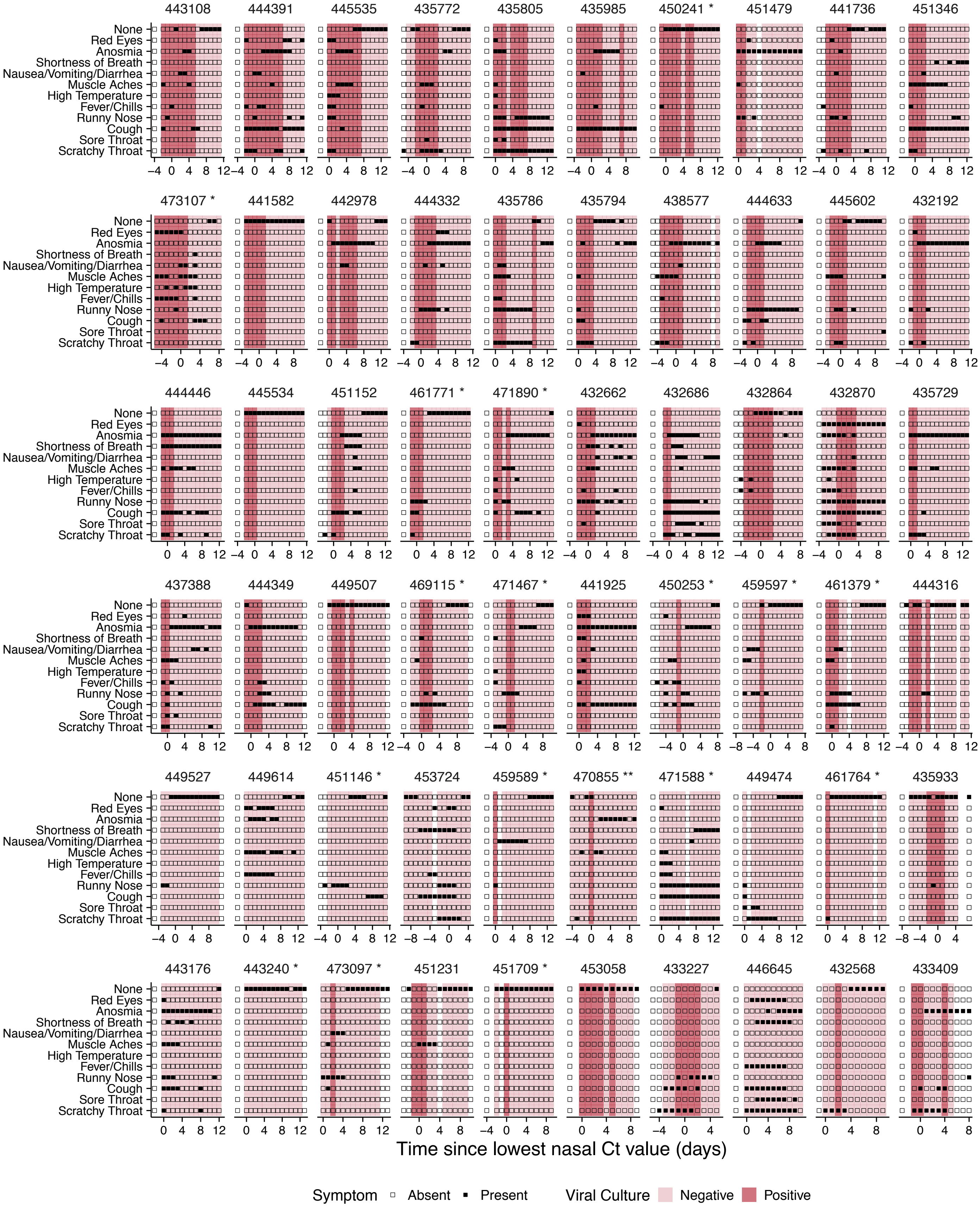


**Fig S2: Individual-level symptom data.** Self-reported symptom data from study participants, overlaid with viral culture status. Participants were asked to complete a survey through the Eureka digital study platform inquiring about the presence or absence of the indicated set of symptoms each day after sample collection.

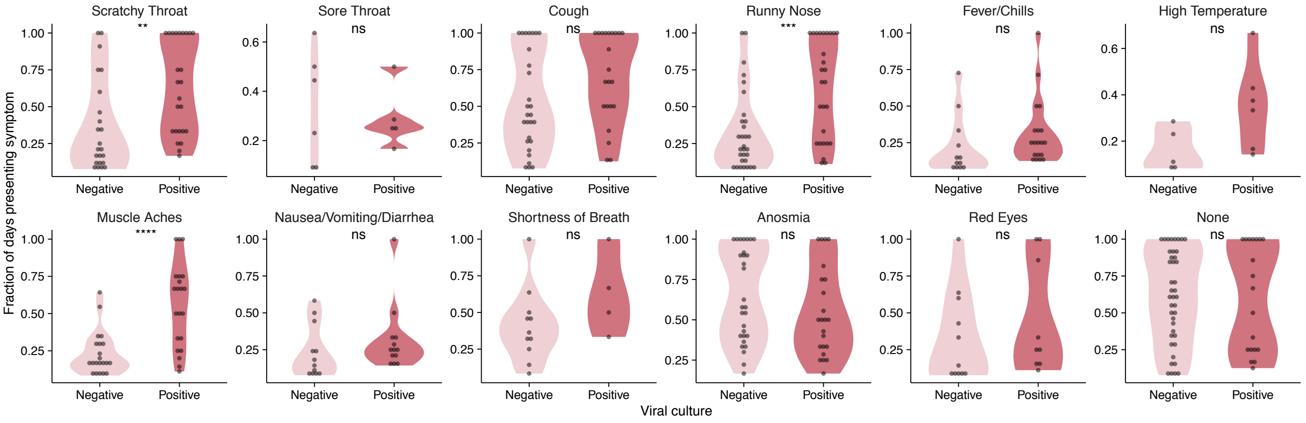


**Fig S3: Comparison of symptoms and viral culture status.** Plots show the proportions of either viral culture negative or viral culture positive days for which participants reported the indicated symptoms. Data are only shown for individuals who reported the indicated symptom at least once.

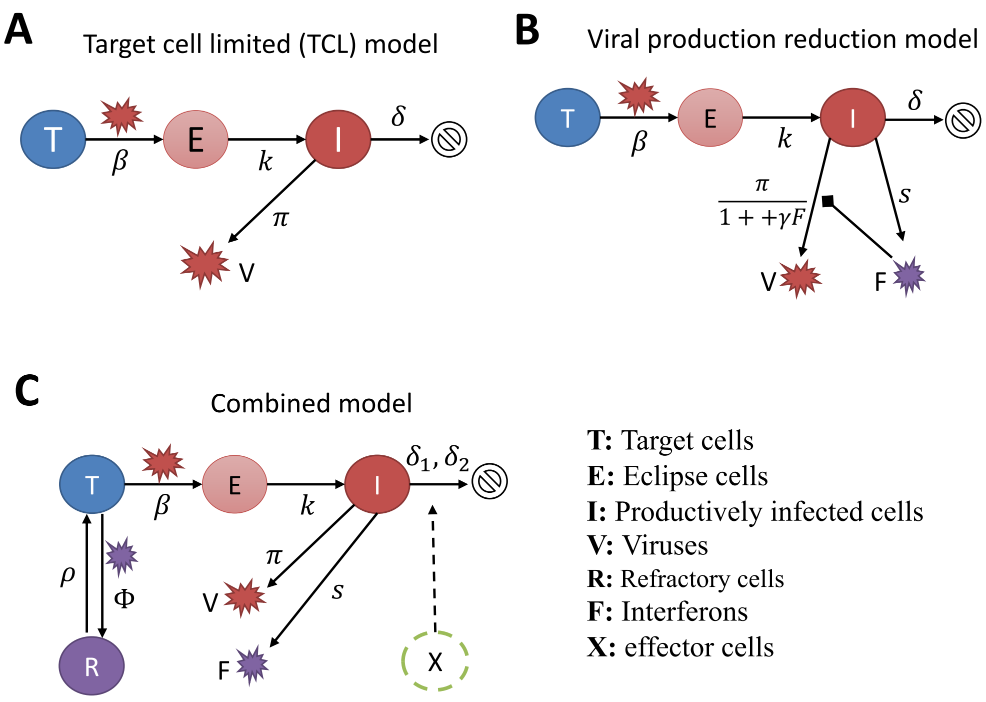


**Fig S4: Model structures.** Diagrams showing the structures of three additional models (not shown in figure 2A) considered for describing viral load data. See Supporting Text for descriptions of the models.

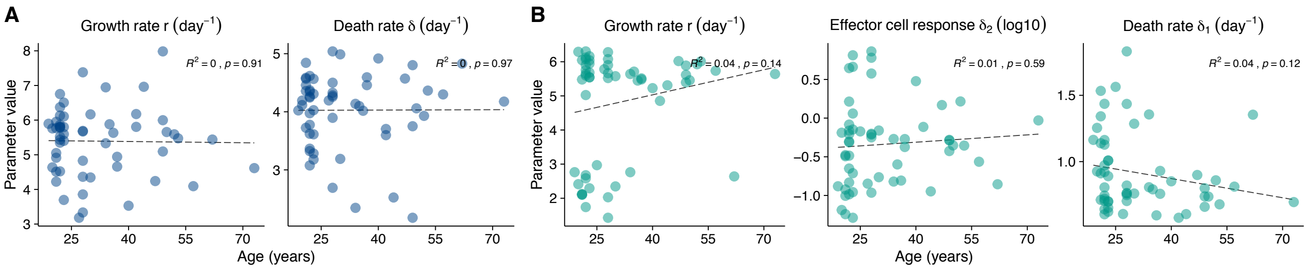


**Fig S5: Model parameter estimates as a function of age.** Plots showing the relationship between age and the indicated model parameter estimates for **(A)** the refractory cell model (nasal data) and the **(B)** the immune effector cell model (saliva data). Linear regressions were performed on the data. $R^2$ values and p-values are shown.

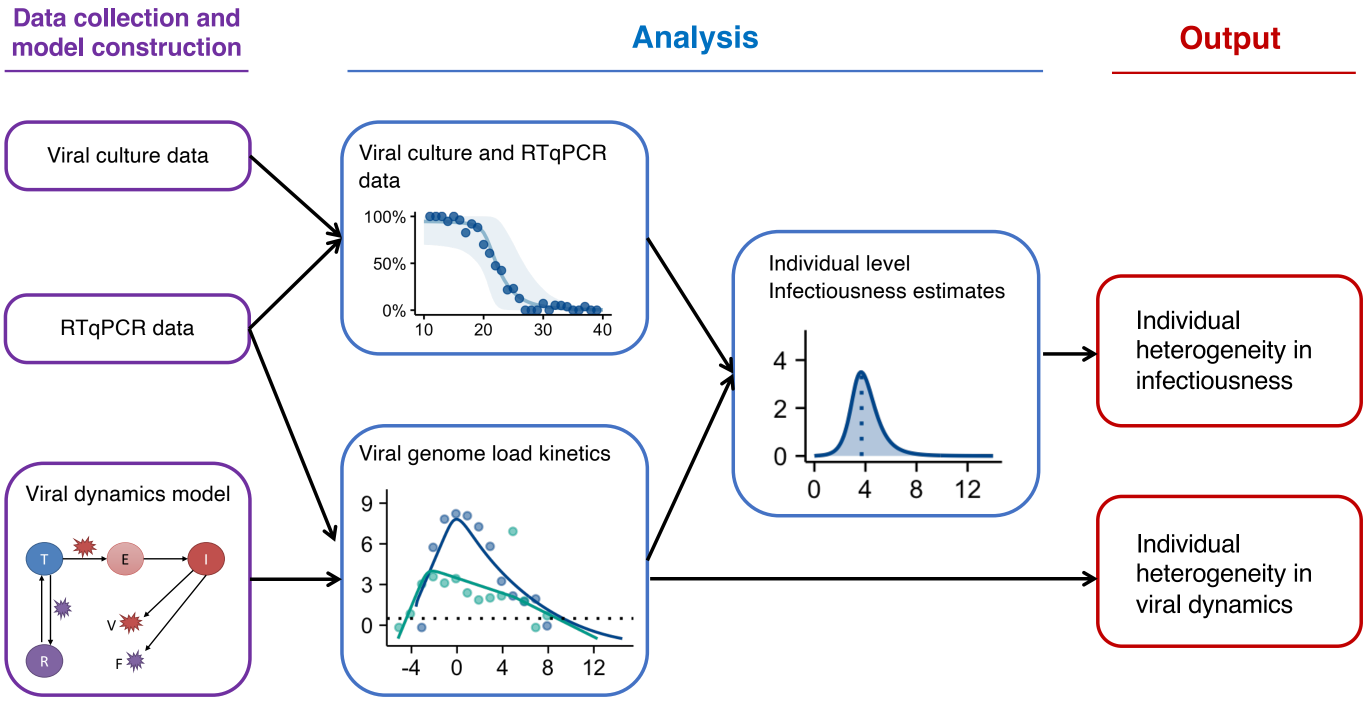


**Fig S6: Analysis workflow.** Diagram indicating how empirical RTqPCR and viral culture data were used to generate estimations of individual level viral dynamics and infectiousness.

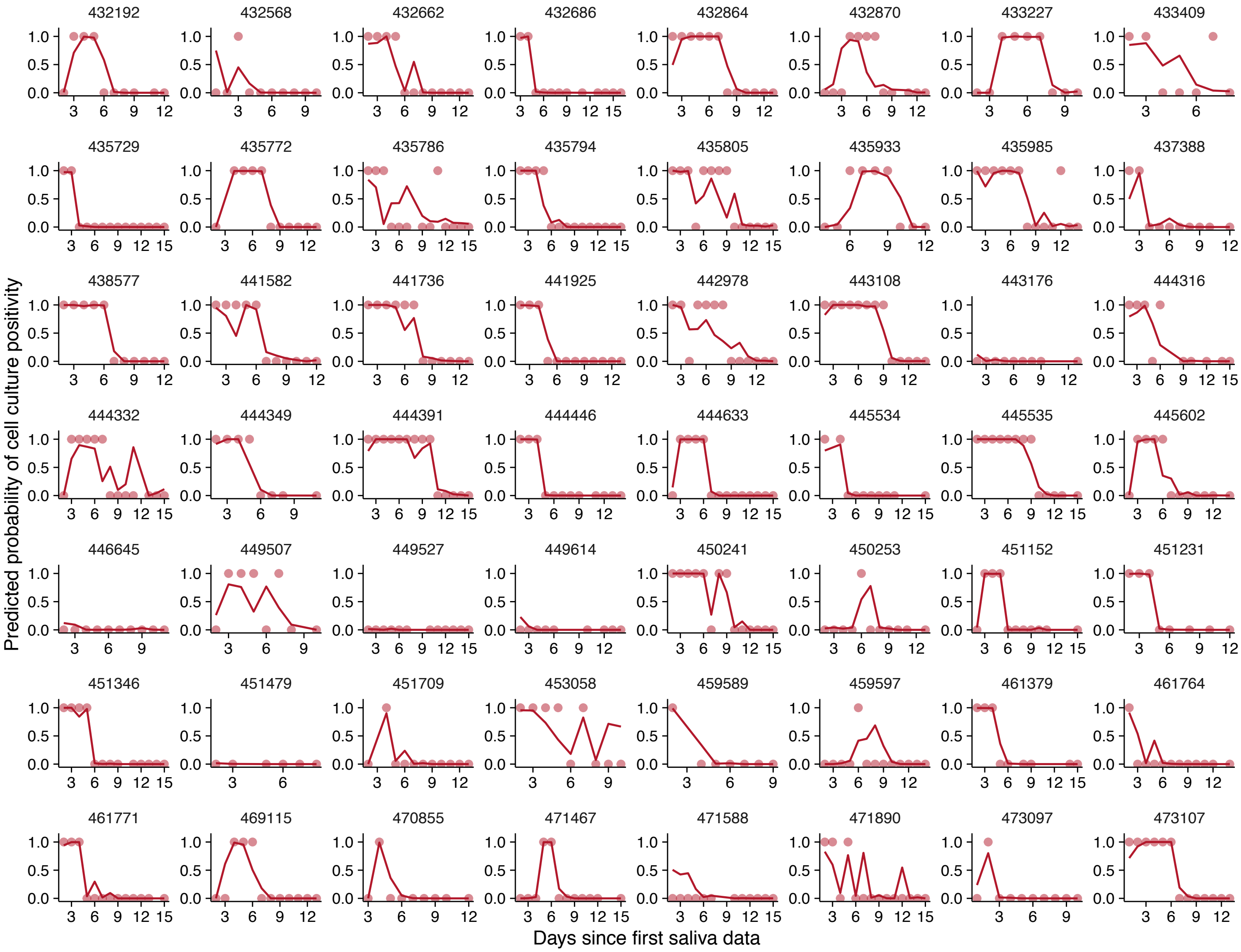


**Fig S7: The saturation model accurately predicts the cell culture positivity data.** Lines denote the predicted probability of cell culture being positive. Dots denotes cell culture positivity data, where a dot is at 1 or 0 when the cell culture is positive or negative, respectively.

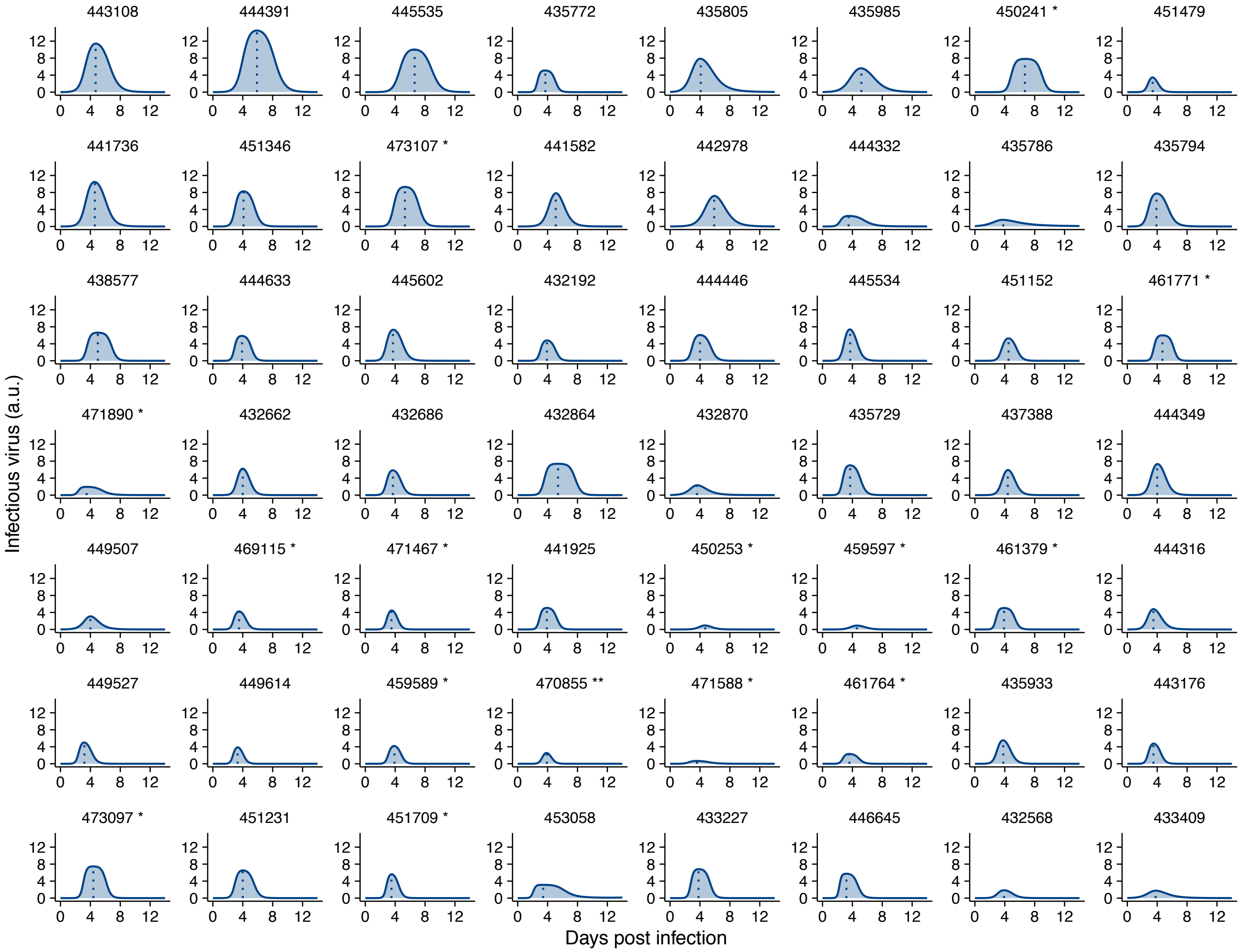


**Fig S8: Individual infectiousness plots.** Estimated infectiousness over time plotted for individual study participants. Dashed lines indicate inferred peak in infectiousness.
