## Supporting Text for "Daily sampling of early SARS-CoV-2 infection reveals substantial heterogeneity in infectiousness"

### 1. Overview of model construction and parameter estimation

The goal of the quantitative analyses is to use mathematical models to characterize viral shedding dynamics from viral genome load data measured by RTqPCR as well as viral culture data that tests the presence of infectious virus. Analyzing the model results, we quantify the individual-level heterogeneity in both viral genome shedding dynamics and infectiousness. See **Fig S6** for an overview of the analysis workflow.

First, we construct viral dynamic models and fit the models to viral genome load data. We estimate key parameters governing the infection process in the nasal and the saliva compartments, such as the viral exponential growth rate before peak viral genome load and the viral clearance rate. This allows us to characterize the individual level heterogeneity in infection kinetics.

Second, we construct mathematical models to describe how the number of infectious virus changes with changes in the viral genome load. We fit the models to viral culture assay data. Using the best model and the predicted viral genome load kinetics from the viral dynamic model, we predict the infectious viral genome load, i.e. the infectiousness, for each individual, and thus quantify the individual-level heterogeneity in infectiousness.

### 2. The viral dynamic models

We constructed viral dynamic models to describe the viral genome load measurements from nasal swabs and saliva samples. The viral genome load patterns in nasal and saliva samples are distinct from each other in many individuals, suggesting compartmentalization of infection dynamics in these two physiological compartments. Therefore, we use the models below to describe data collected from these two compartments separately. See **Fig 2A** and **S4** for schematics of these models.

***The target cell limited (TCL) model***

We first construct a within-host model based on the target cell limited (TCL) model used for other respiratory viruses, such as influenza (*1*) and recently SARS-CoV-2 (*2, 3*). We keep track of the total numbers of target cells (*T*), cells in the eclipse phase of infection (*E*), i.e. infected cells not yet producing virus, productively infected cells (*I*) and viruses (*V*). The ordinary differential equations are:

$$
\begin{aligned}
\frac{dT}{dt} &= -\beta VT \\
\frac{dE}{dt} &= \beta VT - kE \\
\frac{dI}{dt} &= kE - \delta I \qquad \text{[S1]} \\
\frac{dV}{dt} &= \pi I - cV
\end{aligned}
$$

In this model, target cells are infected by virus with rate constant $\beta$. Cells in the eclipse phase become productively infected cells at per capita rate $k$. Productively infected cells die at per capita rate $\delta$. We use *V* to describe viruses measured in nasal or saliva samples and it is a proportion of the total virus in the compartment under consideration. Therefore, the rate, $\pi$, is the product of the viral production rate per infected cell and the proportion of virus that is sampled. See Ke et al. (*3*) for a detailed derivation. Viruses are cleared at per capita rate *c*.

***Refractory cell model***
We extend the TCL model by including an early innate response, i.e. the type-I interferon response, where interferons are produced from infected cells and then bind to receptors on target cells stimulating an antiviral response that makes target cells refractory to viral infection.

We keep track of type I interferon (*F*) and cells refractory to infection (*R*), in addition to other quantities in the TCL model. The full ODEs for target cells, refractory cells and interferon are

$$\frac{dT}{dt} = -\beta VT - \phi FT + \rho R$$
$$\frac{dR}{dt} = \phi FT - \rho R$$
$$\frac{dE}{dt} = \beta VT - kE$$
$$\frac{dI}{dt} = kE - \delta I \qquad \text{[S2]}$$
$$\frac{dV}{dt} = \pi I - cV$$
$$\frac{dF}{dt} = sI - \mu F$$

In this model, the impact of the innate immune response is to convert target cells into refractory cells at rate $\phi FT,$ where $\phi$ is a rate constant. Refractory cells can become target cells again at rate $\rho$. Interferon is produced and cleared at rates $s$ and $\mu$, respectively.

For simplicity and due to a lack of empirical data on interferon responses in our study, we simplify the model by making the quasi-steady-state assumption that the interferon dynamics are much faster than the dynamics of infected cells and assume that $\frac{dF}{dt} = 0$. Thus $sI = \mu F$ or $F = \frac{s}{\mu} I$.

Let $\Phi = \phi \frac{s}{\mu}$ , so that the ODEs for the innate immunity model become:

$$\frac{dT}{dt} = -\beta VT - \Phi IT + \rho R$$
$$\frac{dR}{dt} = \Phi IT - \rho R$$
$$\frac{dE}{dt} = \beta VT - kE \qquad \text{[S3]}$$
$$\frac{dI}{dt} = kE - \delta I$$
$$\frac{dV}{dt} = \pi I - cV$$

***Viral production reduction model***
In addition to making target cells refractory to infection, the impact of interferons may include reducing virus production from infected cells. We include this action of interferons in the viral production reduction model. As above, we make the quasi-steady-state assumption that the interferon dynamics are much faster than the dynamics of infected cells and assume that $F$ is proportional to *I*. The ODEs for the model are

$$\frac{dT}{dt} = -\beta VT \qquad \text{[S4]}$$

$$\frac{dE}{dt} = \beta VT - kE$$
$$\frac{dI}{dt} = kE - \delta I$$
$$\frac{dV}{dt} = \frac{\pi}{1+\gamma I} I - cV$$

where $\gamma$ is a constant representing the effect of interferon in reducing viral production.

***Immune effector cell model***

Over the course of infection, immune effector cells are activated and recruited to kill infected cells. These immune effector cells include innate immune cells such as macrophages and natural killer cells, as well as cells developed during the adaptive immune response, such as cytotoxic T lymphocytes. To consider the impact of these immune effector cells, we develop a model, i.e. the effector cell model, based on a previous model for influenza infection (*4*). In this model, we assume that the death rate of infected cells is $\delta_1$ at the beginning of the infection. This may reflect the cytotoxic effects of viral infection. After time $t_1$, the death rate of infected cells increases by $\delta_2$, where $\delta_2$ models the killing of infected cells by immune effector cells. The ODEs for the model are

$$\frac{dT}{dt} = -\beta VT$$
$$\frac{dE}{dt} = \beta VT - kE$$
$$\frac{dI}{dt} = kE - \delta(t) I \qquad [S5]$$
$$\frac{dV}{dt} = \pi I - cV$$
$$\delta(t) = \begin{cases} \delta_1 & t < t_1 \\ \delta_1 + \delta_2 & t \geq t_1 \end{cases}$$

***Combined model***

In the full model, we combine the refractory cell model and the immune effector cell model to consider both the immediate interferon response and the immune effector response. The ODEs for the model are

$$\frac{dT}{dt} = -\beta VT - \Phi IT + \rho R$$
$$\frac{dR}{dt} = \Phi IT - \rho R$$
$$\frac{dE}{dt} = \beta VT - kE$$
$$\frac{dI}{dt} = kE - \delta(t) I \qquad [S6]$$
$$\frac{dV}{dt} = \pi I - cV$$
$$\delta(t) = \begin{cases} \delta_1 & t < t_1 \\ \delta_1 + \delta_2 & t \geq t_1 \end{cases}$$

***Choice parameter values:***
Total target cell numbers ($T_0$)
We calculate the total numbers of target cells in the nasal and the saliva compartments by multiplying the total number of epithelial cells in these two compartments by the fraction of epithelial cells being targets for SARS-CoV-2 infection.

For the total number of epithelial cells in the nasal compartment, we use the estimate from Baccam et al., i.e. $4\times10^8$ cells (*1*). This is calculated from the estimate that the surface area of nasal turbinates is 160 $cm^2$ (*5*), and the surface area per epithelial cell is $2 \times 10^{-11}$ to $4 \times 10^{-11}$ $m^2$/cell (*1*). For the saliva compartment, the total surface area of the mouth was estimated to be 214.7 $cm^2$ (*6*). Therefore, we estimate that the total number of epithelial cells in the mouth is approximately $4\times10^8\times214.7/160=5.4\times10^8$ cells.

Hou et al. estimated that the fraction of cells that express angiotensin-converting enzyme 2 (ACE2), i.e. the receptor for SARS-CoV-2 entry, on cell surface is approximately 20% in the upper respiratory tract (URT) (*7*). Therefore, in our model, the initial numbers of target cells are calculated as $4\times10^8\times20\%=8\times10^7$ cells and $5.4\times10^8\times20\%=1.08\times10^8$ cells, in the nasal and the saliva compartment, respectively.

Note that these estimates are approximations using available best-estimates in the literature. For a standard viral dynamics model, the number of initial target cells and the virus production rate are unidentifiable and only their product is identifiable (*8*). Thus, if the actual number of target cells differs from what we estimated here, an increase (decrease) in the initial number of target cells will lead to corresponding decrease (increase) in the estimate of the virus production rate.

The initial number of infected cells ($E_0$)
We assume that one cell in the compartment of interest is infected at the start of infection, $E_0 = 1$ cell, consistent with Refs. (*3*) and (*9*). The small number of infected cells is also consistent with a recent work that estimated from sequencing data that the transmission bottleneck is small for SARS-CoV-2 and there are likely 1-3 infected cells at the initiation of infection (*10*). Note that, in an earlier work, we showed that changes in the number of initially infected cells between 1-5 in the model do not substaintially change the inference results (*3*).

***The initial viral growth rate, r***
For all the models, the initial growth of the viral population before peak viral genome load is dominated by viral infection. This means that the immune responses considered in our models act to change the viral growth trajectory substantially only at later time points. Thus, we derive the expression for the initial viral growth rate using the TCL model only (Eqn. S1). This expression represents a good approximation for other models.

At the initial stage of infection, the number of infected cells is orders of magnitude lower than the number of target cells. Thus, it is reasonable to assume the number of target cells is at a constant level, $T_0$. Then, Eqn. S1 becomes a system of linear ODEs, and the initial growth rate *r* is the leading eigenvalue of the Jacobian matrix of the ODE system. We get:

$$r = \frac{1}{2}\left[-(k+\delta) + \sqrt{(k+\delta)^2 + 4k\delta(R_0 - 1)}\right] \quad \text{(S7)}$$

where $R_0 = \frac{\beta\pi}{\delta c} T_0$.

***Model fitting strategy***
Fitting viral dynamic models to the viral genome load data

We took a non-linear mixed effect modeling approach to fit the viral dynamic models to viral genome load data from all individuals simultaneously. All estimations were performed using Monolix (Monolix Suite 2019R2, Antony, France: Lixoft SAS, 2019. lixoft.com/products/monolix/). We allowed random effects on the fitted parameters (unless specified otherwise). All population parameters except for the starting time of simulation, $t_0$, are positive and therefore, we assume that they follow log-normal distributions. For $t_0$, we assume a normal distribution because $t_0$ can be positive or negative.

The parameters $\beta$ and $\pi$ in the viral dynamic models strongly correlate with each other when the models are fitted to viral genome load data (*8*). We tested three choices in handling this correlation in fitting all 5 viral dynamic models: 1) a correlation is assumed between parameter $\beta$ and $\pi$ in Monolix, 2) parameter $\beta$ has fixed effect only (i.e. its value is set to be the same across all individuals), and 3) parameter $\pi$ has fixed effect only.

To test if the age of the individuals and the type of viruses infecting an individual (grouped in two as non-B.1.1.7 and B.1.1.7) explains the heterogenous patterns in the viral genome load trajectories across the cohort, we test whether they covary with any of the fitted parameters in the model by setting the two variables as a continuous and a categorical covariate, respectively, of the in Monolix.

The assumptions on parameters $\beta$ and $\pi$ and the choice of parameters that covariate with age or viral strain of infection lead to a large number of model choices for fitting. Therefore, we took the following strategy to ensure we identify the best model and parameter combinations to describe the data:

- First, we test the three assumptions about parameters $\beta$ and $\pi$ in the five viral dynamic models without any covariate, and select the best assumption for further analysis based on their AIC scores.
- Second, using the best assumption, we test the model by including the age of the individuals as a continuous covariate of all fitted parameter values with a random effect first. We then take an iterative approach to test whether if the covariate should be removed from any of the parameters in the model using the Pearson's correlation test in Monolix. The parameter(s) that has a non-significant p-value (p-value>0.05) or with the lowest p-value is removed from next round of parameter fitting. We iterate the process until all parameters are removed.
- Then, the best model variant with the lowest AIC score is selected for analysis on whether parameter estimates differ in individuals infected by different viral strains. As before, we take an iterative approach. We first set the viral strain, i.e. non-B.1.1.7 or B.1.1.7 as a categorical covariate of all fitted parameter values with a random effect in the model. We then test whether if the covariate should be removed from any of the parameters in the model using the Analysis of Variance (ANOVA) in Monolix. The parameter(s) that has a non-significant p-value (p-value>0.05) or with the lowest p-value is removed from next round of parameter fitting. We iterate the process until all parameters are removed.
- Finally, the model variant with the lowest AIC score is selected as the best model.

***Predicting viral genome load trajectories for non-B1.1.7 and B1.1.7 strains***

We randomly sample 5000 sets of parameter combinations from the distribution specified by the best-fit population parameters (Table S4). For the effector cell model for the saliva compartment, $\beta$ and $\pi$ are strongly correlated. We thus applied formulations such that the correlations between the two parameter values are preserved in the random sampling in accordance with the estimated correlation coefficient. We simulate the best-fit model using the 5000 sets of parameter combinations for each of the strain. The median and the $5^{th}$ and $95^{th}$ quantile of viral genome loads at each time points are reported.

### 3. Viral genome load calibration from Ct/Cn values and their impacts on estimates of key parameters in viral dynamic models

***Viral genome load calibration***

To calculate viral genome loads from Ct/Cn values, we first performed experiments on 4 nasal samples to measure the Cn values and quantify their viral genome load. For each sample, measurements were performed on both N1 and N2 genes. For the sample with a Cn value of 17.48, three replicate experiments were performed. See Table A1 below for the measured Cn values and their corresponding viral genome loads. We then performed a linear regression on the measured Cn values and the $\log_{10}$ viral genome loads. This leads to the following formular for the relationship between Cn values and viral genome load:

$$\log_{10} V = 13.02 - 0.33Cn \qquad [S8]$$

where $V$ and $Cn$ denote the viral genome load and the Cn value, respectively. Note that Eqn. S8 can also be expressed as $V = 1.05 \times 10^{13} \times 2.1^{-Cn}$, i.e. an increase of Cn by 1 translates to approximately 2 fold decrease in viral load.

**Table A1. Cn values and the measured viral genome loads used for the viral genome load calibration curve.**

| Cn value | Viral genome load (/μL) | Gene |
|---|---|---|
| 21.95 | 588 | N1 |
| 26.02 | 12.7 | N1 |
| 29.8 | 2.52 | N1 |
| 17.48 | 19832.5 | N1 |
| 17.48 | 20720.1 | N1 |
| 17.48 | 20280.2 | N1 |
| 21.95 | 579.0 | N2 |
| 26.02 | 12.1 | N2 |
| 29.8 | 3.05 | N2 |
| 17.48 | 18947.7 | N2 |
| 17.48 | 19542.4 | N2 |
| 17.48 | 19288.4 | N2 |

The RTqPCR used to measure Ct values from saliva samples has similar efficiency as the RTqPRC used to measure Cn values from nasal samples; however, the calibration curve for Ct values measured from saliva sample is not available to us. Therefore, we used the same formulation (i.e. Eqn. S8) to translate Ct values to viral genome loads in saliva. Because of the similar efficiency between the two RTqPCR platforms, we expect the coefficient -0.33 in Eqn. S8 represents an accurate description of how viral genome load changes with changes in Ct values for the RTqPCR used for saliva samples. However, as mentioned in the main text, saliva RTqPCR assay does not include an RNA extraction step, leading to a lower overall sensitivity for measuring viral RNAs from saliva samples. Therefore, we expect that the predicted viral genome load using Eqn. S8 is a constant (unknown) fraction of the actual viral genome load. In mathematical terms, we expect the first constant 13.02 in Eqn.8 to be larger for saliva samples.

***Impacts on estimates of key parameters in viral dynamic models***
To understand the impact of using calibrated viral loads that are a constant proportion to the true viral loads in model inference, here we analyze how it affects parameter estimates in viral dynamic models. We let the actual viral genome load be $V$, and the viral genome load predicted by Eqn. S8 be $V^*$, and $V^* = aV$.

We first use the ODEs for the TCL model (i.e. Eqn. S1) as an example. Substituting $V^* = aV$ into the ODEs in Eqn. S1, we get:

$$\begin{aligned}\frac{dT}{dt} &= -\frac{\beta}{a}V^*T \\ \frac{dE}{dt} &= \frac{\beta}{a}V^*T - kE \\ \frac{dI}{dt} &= kE - \delta I \\ \frac{dV^*}{dt} &= a\pi I - cV^*\end{aligned} \qquad [S9]$$

Comparing Eqn. S9 with S1, we note that the only differences are that parameters $\beta$ and $\pi$ in Eqn. S1 are scaled by $\frac{1}{a}$ and $a$, respectively in Eqn. S9.

The analysis above shows that when we use viral genome load data that are a constant proportion of the true viral genome load, the only differences are that the estimated values of $\beta$ and $\pi$ are scaled by the proportion. Estimates of other parameter values in the model as well as the exponential growth rate, r (Eqn. S7) are unchanged. Note that the scaling on $\beta$ and $\pi$ are cancelled out in the expression of $R_0$ in Eqn. S7.

Extending this observation to other viral dynamic models we used, we expect that
- the parameter $\Phi$ in refractory cell model for the saliva compartment has a scaling factor of $\frac{1}{a}$, similar as $\beta$.
- Parameters in the models (other than $\beta$, $\pi$ and $\Phi$) are not affected by the scaling.

The scaling also explains the fact that the estimated values of $\beta$ or $\pi$ differ orders of magnitude between the two models for the nasal and saliva compartments (see **Table S4 and S5**). Therefore, this difference in estimates of $\beta$ or $\pi$ between the two compartment largely arises from the lower molecular sensitivity of the saliva assay than the nasal assay.

### 4. Modeling infectiousness of an individual
We model how infectiousness depends on the viral genome load in an individual similar to the framework proposed in Ke et al. (*11*). Specifically, we first use the viral culture data collected in this study to infer how the level of infectious viruses depends on viral genome load. From this model, we predict how the level of infectious viruses changes over time in each individual and how the overall infectiousness of the infection varies among the participants.

***Relationship between viral genome load and infectious viruses***
We first consider three alternative models describing how the number of infectious viruses in a sample is related to viral genome load measured by RTqPCR: the 'linear' model, 'power-law' model and the 'saturation' model. In these models, due to the nature of stochasticity in sampling, we assume the number of infectious viruses that was in the sample for cell culture experiment to

be a random variable, *Y*, that follows a Poisson distribution, and $V_{inf}$ represents the expected number of infectious viruses, i.e. $V_{inf} = E(Y)$.

1. The linear model

We assume that $V_{inf}$, is proportional to the viral genome load, *V*, in the sample:

$$V_{inf} = E(Y) = AV \qquad \text{[S10]}$$

where *A* is a constant.

2. The power-law model

We assume that $V_{inf}$, is related to the viral genome load, *V*, by a power function:

$$V_{inf} = E(Y) = BV^h \qquad \text{[S11]}$$

where *B* and *h* are constants.

3. The saturation model

We assume that $V_{inf}$, is related to the viral genome load, *V*, by a Hill function

$$V_{inf} = E(Y) = V_m \frac{V^h}{V^h + K_m^h} \qquad \text{[S12]}$$

where $V_m$ and $K_m$ are constants and *h* is the Hill-coefficient.

***Probability of cell culture being positive***

If each infectious virus has a probability $\varrho$ to establish infection such that the cell culture becomes positive, the number of viruses that successfully establish an infection in cell culture is Poisson distributed with parameter $\lambda = E(Y)\varrho = V_{inf}\varrho$. Thus, the probability of one or more viruses successfully infecting the culture so that it tests positive is

$$p_{positive} = 1 - \exp(-\lambda) = 1 - \exp\,(-V_{inf}\varrho) \qquad \text{[S13]}$$

Substituting the expressions of $V_{inf}$ from the three models above, we get the following expressions for $p_{positive}$ from the three models (note that we use the subscripts '1', '2' and '3' to denote the three models for $V_{inf}$):

$$p_{positive,1} = 1 - \exp\left(-V_{inf}\varrho\right) = 1 - \exp(-DV) \qquad \text{[S14]}$$

where $D = A\varrho$.

$$p_{positive,2} = 1 - \exp\left(-V_{inf}\varrho\right) = 1 - \exp\left(-GV^h\right) \qquad \text{[S15]}$$

where $G = B\varrho$.

$$p_{positive,3} = 1 - \exp\left(-V_{inf}\varrho\right) = 1 - \exp\left(-J\frac{V^h}{V^h + K_m^h}\right) \qquad \text{[S16]}$$

where $J = V_m\varrho$.

Note that from the expressions above, it becomes clear that we will not be able to estimate parameters A, B and $V_m$ in the three models, because they appear as products with the unknown parameter $\varrho$ in the equations. This means that the viral culture data does not allow us to estimate the absolute number of infectious viruses in a sample or given a viral genome load; instead, we are able to estimate a quantity that is a constant proportion of the actual number of infectious viruses over time and across individuals. Therefore, we report estimations of infectious viruses in an arbitrary unit (a.u.). These estimates represent relative measure of infectiousness. Two estimates measured at different time points and/or from different individuals can be compared with each other.

***Model fitting using a population effect modeling approach***

For each sample, the viral genome load and the cell culture positivity were measured. Using this data, we estimate parameter values in the three models by minimizing the negative log-likelihood of the data.

More specifically, the likelihood of the m$^{th}$ observation being positive or negative in cell culture is calculated as

$$p_{i,m} = \begin{cases} p_{positive,i}(V_m), & if\ the\ kth\ observation\ is\ positive \\ 1 - p_{positive,i}(V_m), & if\ the\ kth\ observation\ is\ negative \end{cases} \qquad [S17]$$

where $V_m$ is the viral genome load of the $m^{th}$ observation.

Because we have the paired nasal RTqPCR and cell-culture positivity data for each individual, we fit the three mathematical models using a non-linear mixed effect modeling approach. Again, all estimations were performed using Monolix (Monolix Suite 2019R2, Antony, France: Lixoft SAS, 2019. lixoft.com/products/monolix/). We allowed random effects on the fitted parameters (unless specified otherwise). All population parameters with a random effect are assumed to follow log-normal distributions.

To find the best-model explaining the data, we tested models with different combination of parameters with either a random effect or without a random effect (**Table S7**). The model with the lowest AIC score is selected as the best model.

Note that for each of the three models, we tested a model variation where all parameters in the models have fixed effects only, i.e. a single set of parameters is used to explain viral culture data from every individual. In this case, there is no heterogeneity in parameter values across individuals. The resulting AIC scores are significantly worse than the best-fit model assuming random effects on parameters (see **Table S7**). This indicates that there exists substantial level of individual heterogeneity in the relationship between infectious viruses shedding and viral genome loads (as shown in **Fig 3D**).

***Calculating confidence intervals of the cell-culture-positivity curve (in Fig. 3C)***

Similar as the procedures performed for predicting the confidence intervals of viral genome load trajectories, we randomly sample 5000 sets of parameter combinations from the distribution specified by the best-fit population parameters of the best model, i.e. the saturation model assuming $K_m$ only has a fixed effect (**Table S8**). More specifically, we sampled parameters from a log-normal distribution for $J$ and $h$, with their means and standard deviations at the best-fit values. We set $K_m$=4x10$^6$ /mL, i.e. the best-fit parameter for all combinations. The Using the parameter combinations, we generated curves of probability of cell-culture positivity at Cn values ranging between 10 and 40. The median and the 5$^{th}$ and 95$^{th}$ quantile of viral genome loads at each Cn value are reported.
