## Supplemental Tables for "Daily sampling of early SARS-CoV-2 infection reveals substantial heterogeneity in infectiousness"

**Table S1: Study cohort demographics information**

| | |
|---|---|
| **N** | 60 |
| **age (median [range])** | 28.00 [19.00, 73.00] |

| | | |
|---|---|---|
| **race (%)** | Asian | 2 (3.3) |
| | Black | 6 (10.0) |
| | Other | 6 (10.0) |
| | White | 46 (76.7) |
| **gender (%)** | Female | 26 (43.3) |
| | Male | 34 (56.7) |
| **ethnicity (%)** | Hispanic | 10 (16.7) |
| | Non-Hispanic | 50 (83.3) |

| **Self-reported conditions:** | | |
|---|---|---|
| **copd (%)** | No | 59 (98.3) |
| | Yes | 1 (1.7) |
| **asthma (%)** | No | 58 (96.7) |
| | Yes | 2 (3.3) |
| **cancer (%)** | No | 58 (96.7) |
| | Yes | 2 (3.3) |
| **immunodeficiency (%)** | No | 58 (96.7) |
| | Yes | 1 (1.7) |
| | NA | 1 (1.7) |
| **anemia (%)** | No | 58 (96.7) |
| | Yes | 1 (1.7) |
| | NA | 1 (1.7) |

**Table S2. Comparing model fits to longitudinal viral load data from nasal swab samples using AIC scores.** The best overall model is the refractory cell model assuming the innate immune parameter $\Phi$ covaries with the age of individuals (its AIC score is bolded and underlined).

| Model | Assumptions on $\beta$ and $\pi$ | Covariate | | AIC |
|---|---|---|---|---|
| | | Age | Viral type | |
| TCL model | Correlation | - | - | 2256.9 |
| | Fixed effect $\beta$ | - | - | 2365.2 |
| | Fixed effect $\pi$ | - | - | 2364.7 |
| | Correlation | $t_0, \beta, \delta, \pi$ | - | 2257.4 |
| | Correlation | $t_0, \beta, \pi$ | - | 2259.7 |
| | Correlation | $\beta, \pi$ | - | 2254.7 |
| | Correlation | $\beta$ | - | 2254.5 |
| | Correlation | - | $t_0, \beta, \delta, \pi$ | 2282.7 |
| | Correlation | - | $t_0, \beta$ | 2299.1 |
| | Correlation | - | $\beta$ | 2252.1 |
| Refractory cell model | Correlation | - | - | 2187.9 |
| | Fixed effect $\beta$ | - | - | 2181.4 |
| | Fixed effect $\pi$ | - | - | 2182.4 |
| | Fixed effect $\beta$ | $t_0, \delta, \pi, \Phi, \rho$ | - | 2184.7 |
| | Fixed effect $\beta$ | $\Phi, \rho$ | - | 2180 |
| | Fixed effect $\beta$ | $\Phi$ | - | **2176.7** |
| | Fixed effect $\beta$ | $\Phi$ | $t_0, \delta, \pi, \Phi, \rho$ | 2182 |
| | Fixed effect $\beta$ | $\Phi$ | $\delta, \pi, \rho$ | 2177.4 |
| | Fixed effect $\beta$ | $\Phi$ | $\pi, \rho$ | 2178.8 |
| | Fixed effect $\beta$ | $\Phi$ | $\pi$ | 2178.7 |
| Viral production reduction model | Correlation | - | - | 2263.2 |
| | Fixed effect $\beta$ | - | - | 2270 |
| | Fixed effect $\pi$ | - | - | 2265 |
| | Correlation | $t_0, \beta, \delta, \pi, \gamma$ | - | 2262 |
| | Correlation | $\beta, \pi, \gamma$ | - | 2255.2 |
| | Correlation | $\beta, \pi$ | - | 2258 |
| | Correlation | $\pi$ | - | 2264.2 |
| | Correlation | $\beta, \pi, \gamma$ | $t_0, \beta, \delta, \pi, \gamma$ | 2272 |
| | Correlation | $\beta, \pi, \gamma$ | $\beta, \pi, \gamma$ | 2258.6 |
| Immune effector cell model | Correlation | - | - | 2258.6 |
| | Fixed effect $\beta$ | - | - | 2271.5 |
| | Fixed effect $\pi$ | - | - | 2271.3 |
| | Correlation | $t_0, \beta, \delta_1, \pi, t_1, \delta_2$ | - | 2268.1 |
| | Correlation | $\beta, \pi, \delta_2$ | - | 2266.6 |
| | Correlation | $\pi, \delta_2$ | - | 2275.6 |
| | Correlation | $\delta_2$ | - | 2266.8 |
| | Correlation | - | $t_0, \beta, \delta_1, \pi, t_1, \delta_2$ | 2271.1 |
| | Correlation | - | $\delta_1, t_1$ | 2269.7 |

| | | | | |
|---|---|---|---|---|
| | Correlation | - | $t_1$ | 2259.1 |
| Combined model | Correlation | - | - | 2196.7 |
| | Fixed effect $\beta$ | - | - | 2190.1 |
| | Fixed effect $\pi$ | - | - | 2188.5 |
| | Fixed effect $\pi$ | $t_0, \beta, \delta_1, \Phi, \rho, t_1, \delta_2$ | - | 2198.1 |
| | Fixed effect $\pi$ | $\delta_1, \Phi, \rho, t_1, \delta_2$ | - | 2190.4 |
| | Fixed effect $\pi$ | $\Phi, \rho, t_1, \delta_2$ | - | 2189.4 |
| | Fixed effect $\pi$ | $\Phi, t_1, \delta_2$ | - | 2189.1 |
| | Fixed effect $\pi$ | $t_1, \delta_2$ | - | 2193.6 |
| | Fixed effect $\pi$ | $t_1$ | - | 2190.9 |
| | Fixed effect $\pi$ | - | $t_0, \beta, \delta_1, \Phi, \rho, t_1, \delta_2$ | 2199.6 |
| | Fixed effect $\pi$ | - | $\beta, \delta_1, \rho, t_1, \delta_2$ | 2194.4 |
| | Fixed effect $\pi$ | - | $\beta, \delta_1, \rho, t_1$ | 2195.8 |
| | Fixed effect $\pi$ | - | $\beta, \rho, t_1$ | 2194.6 |
| | Fixed effect $\pi$ | - | $\rho, t_1$ | 2192.6 |
| | Fixed effect $\pi$ | - | $\rho$ | 2188.9 |

**Table S3. Comparing model fits to longitudinal viral load data from saliva samples using AIC scores.** The best overall model is the effector cell model assuming all fitted parameters covary with viral type, i.e. non-B1.1.7 vs. B1.1.7 (its AIC score is bolded and underlined).

| Model | Assumptions on $\beta$ and $\pi$ | Covariate | | AIC |
|---|---|---|---|---|
| | | Age | Viral type | |
| TCL model | Correlation | - | - | 2296.8 |
| | Fixed effect - $\beta$ | - | - | 2380.2 |
| | Fixed effect - $\pi$ | - | - | 2377.8 |
| | Correlation | $t_0, \beta, \delta, \pi$ | - | 2307.2 |
| | Correlation | - | $t_0, \beta, \delta, \pi$ | 2288.4 |
| | Correlation | - | $t_0, \beta, \delta$ | 2292.6 |
| | Correlation | - | $t_0, \delta$ | 2303.4 |
| | Correlation | - | $t_0$ | 2305.7 |
| | Correlation | - | $\delta$ | 2293.1 |
| Refractory cell model | Correlation | - | - | 2308.8 |
| | Fixed effect $\beta$ | - | - | 2314.7 |
| | Fixed effect $\pi$ | - | - | 2322.4 |
| | Correlation | - | $t_0, \beta, \delta, \pi, \Phi, \rho$ | 2329 |
| | Correlation | - | $t_0, \beta, \delta, \pi, \rho$ | 2333.7 |
| | Correlation | - | $t_0, \beta, \pi, \rho$ | 2324 |
| | Correlation | - | $\beta, \pi, \rho$ | 2318.8 |
| | Correlation | - | $\beta, \rho$ | 2310.9 |
| | Correlation | $t_0, \beta, \delta, \pi, \Phi, \rho$ | - | 2305.7 |
| | Correlation | $\beta, \pi$ | - | 2327.7 |
| | Correlation | $\beta$ | - | 2317.1 |
| | Correlation | $\pi$ | - | 2326.8 |
| Viral production reduction model | Correlation | - | - | 2290.1 |
| | Fixed effect $\beta$ | - | - | 2333.3 |
| | Fixed effect $\pi$ | - | - | 2379.1 |
| | Correlation | $t_0, \beta, \delta, \pi, \gamma$ | - | 2314.6 |
| | Correlation | $\delta, \pi, \gamma$ | - | 2298.9 |
| | Correlation | $\delta, \gamma$ | - | 2297.9 |
| | Correlation | $\delta$ | - | 2301.5 |
| | Correlation | - | $t_0, \beta, \delta, \pi, \gamma$ | 2293.6 |
| | Correlation | - | $t_0, \beta, \delta, \gamma$ | 2306.5 |
| | Correlation | - | $t_0, \delta, \gamma$ | 2292.1 |
| | Correlation | - | $\gamma$ | 2288.1 |
| Immune effector cell model | Correlation | - | - | 2286.5 |
| | Fixed effect $\beta$ | - | - | 2313.8 |
| | Fixed effect $\pi$ | - | - | 2345.3 |
| | Correlation | $t_0, \beta, \delta_1, \pi, t_1, \delta_2$ | - | 2291.4 |
| | Correlation | $t_0$ | - | 2288.9 |
| | Correlation | - | $t_0, \beta, \delta_1, \pi, t_1, \delta_2$ | **<u>2269.4</u>** |
| | Correlation | - | $t_0, \beta, \delta_1, \pi, t_1$ | 2272 |

| | | | | |
|---|---|---|---|---|
| | Correlation | - | $t_0, \beta, \delta_1, t_1$ | 2274 |
| | Correlation | - | $t_0, \delta_1, t_1$ | 2272.6 |
| | Correlation | - | $\beta, \delta_1, \pi$ | 2274 |
| | Correlation | - | $\delta_1$ | 2276.2 |
| Combined model | Correlation | - | - | 2292.4 |
| | Fixed effect $\beta$ | - | - | 2283.7 |
| | Fixed effect $\pi$ | - | - | 2288.2 |
| | Fixed effect $\beta$ | $t_0, \beta, \delta_1, \pi, \Phi, \rho, t_1, \delta_2$ | - | 2294.6 |
| | Fixed effect $\beta$ | $\pi$ | - | 2281.2 |
| | Fixed effect $\beta$ | $\pi$ | $t_0, \beta, \delta_1, \pi, \Phi, \rho, t_1, \delta_2$ | 2282.2 |
| | Fixed effect $\beta$ | $\pi$ | $t_0, \delta_1, \pi, \Phi, \rho, t_1$ | 2276.8 |
| | Fixed effect $\beta$ | $\pi$ | $t_0, \delta_1, \pi, \Phi$ | 2274.4 |
| | Fixed effect $\beta$ | $\pi$ | $t_0, \delta_1, \Phi$ | 2282.7 |
| | Fixed effect $\beta$ | $\pi$ | $t_0, \delta_1$ | 2292.8 |

**Table S4. Estimated population parameter values from the best models, i.e., the refractory cell model for the nasal compartment and the effector cell model for the saliva compartment.** The means and standard deviations are derived assuming that individual parameters follow log-normal distributions.

| Model | Parameter | Description | Mean (population estimate) | Standard deviation* |
|---|---|---|---|---|
| Refractory cell model (Nasal compartment) | $t_0$ | Estimated time of infection in nasal relative to the day of diagnosis | -1.4 days | 1.9 days |
| | $\beta$ | Infectivity parameter constant | $1.69 \times 10^{-9}$ mL/day | - |
| | $\delta$ | Death rate of infected cells | 4.0 /day | 0.27 |
| | $\pi$ | Composite parameter for virus production and sampling | $1.64 \times 10^{3}$ /day | 0.24 |
| | $\Phi$ | Rate constant for the interferon-induced conversion of target cells to refractory cells | $1.43 \times 10^{-4}$ mL/day | 1.37 |
| | $\sigma_{\Phi}$ | Coefficient of the covariate, age, on $\Phi$ | -0.066 | |
| | $\rho$ | Rate at which refractory cells become target cells again | 0.0044 /day | 0.12 |
| Effector cell model (Saliva compartment) | $t_0$ (non-B.1.1.7) | Estimated time of infection in nasal relative to the day of diagnosis | -4.1 days | 1.2 days |
| | $t_0$ (non-B.1.1.7) | | -8.1 days | |
| | $\beta$ (non-B.1.1.7) | Infectivity parameter constant | $5.88 \times 10^{-5}$ mL/day | 2.0 |
| | $\beta$ (B.1.1.7) | | $5.78 \times 10^{-6}$ mL/day | |
| | $\delta_1$(non-B.1.1.7) | Death rate of infected cells in the absence of effector cell responses | 0.75 /day | 0.23 |
| | $\delta_1$(B.1.1.7) | | 1.28 /day | |
| | $\pi$ (non-B.1.1.7) | Composite parameter for virus production and sampling | 0.025 /day | 1.9 |
| | $\pi$ (B.1.1.7) | | 0.094 /day | |
| | $t_1$ (non-B.1.1.7) | Time when the effector cell response is developed | 10.2 day | 0.2 |
| | $t_1$ (B.1.1.7) | | 8.1 day | |
| | $\delta_2$ (non-B.1.1.7) | The increase in infected cell killing due to the effector cell response | 0.58 /day | 1.96 |
| | $\delta_2$ (B.1.1.7) | | 0.17 /day | |

* All the population parameters except for $t_0$ are assumed to follow log-normal distributions in the population, and therefore, the report standard deviation is for a log-normal distribution. Parameter $t_0$ is assumed to be normally distributed.

**Table S5. Individual parameter values estimated from the best fit models, i.e. the refractory cell model and the effector cell model for viral dynamics in the nasal and saliva compartment respectively.** 'NA' denotes individuals where data are not included in parameter estimation.

| ID | Refectory cell model | | | | | | Effector cell model | | | | | |
|---|---|---|---|---|---|---|---|---|---|---|---|---|
| | $t_0$ (day) | $\beta$ ($10^{-9}$ mL/day) | $\delta$ (/day) | $\pi$ (/day) | $\Phi$ ($10^{-6}$ /day) | $\rho$ (/day) | $t_0$ (day) | $\beta$ ($10^{-5}$ mL/day) | $\delta_1$ (/day) | $\pi$ (/day) | $t_1$ (day) | $\delta_2$ (/day) |
| **432192** | -0.1 | 1.69 | 4 | 1642 | 25.2 | 0.011 | 4 | 68.6 | 0.7 | 0.002 | 10.4 | 0.1 |
| **432568** | 3.5 | 1.69 | 4.1 | 1635 | 34.3 | 0.01 | 3.9 | 4.8 | 0.8 | 0.03 | 10.2 | 0.5 |
| **432662** | 0.7 | 1.69 | 4.6 | 1613 | 47.1 | 0.01 | 2.8 | 3.8 | 0.7 | 0.036 | 9.7 | 0.3 |
| **432686** | 1.3 | 1.69 | 3.6 | 1706 | 32.3 | 0.011 | 3.4 | 2.8 | 0.6 | 0.048 | 10.8 | 0.1 |
| **432864** | 0.7 | 1.69 | 4.2 | 1401 | 0.8 | 0.011 | 4.7 | 2.2 | 0.7 | 0.065 | 8.1 | 0.9 |
| **432870** | -0.3 | 1.69 | 4.3 | 1689 | 87.9 | 0.01 | 3.2 | 4.2 | 0.7 | 0.034 | 9.3 | 0.7 |
| **433227** | -1.5 | 1.69 | 3.8 | 1803 | 10.7 | 0.011 | 3.5 | 3.5 | 0.7 | 0.039 | 10.1 | 0.5 |
| **433409** | 1.6 | 1.69 | 4.4 | 1743 | 34.7 | 0.01 | 4.2 | 3.6 | 0.7 | 0.039 | 10.2 | 0.6 |
| **435729** | 2.9 | 1.69 | 3.6 | 1840 | 15.4 | 0.011 | NA | NA | NA | NA | NA | NA |
| **435772** | -1.4 | 1.69 | 4.6 | 2108 | 13.4 | 0.01 | 1.9 | 109 | 0.6 | 0.001 | 10 | 0.3 |
| **435786** | 2.4 | 1.69 | 3.2 | 1760 | 11.1 | 0.011 | 4.2 | 2.7 | 0.8 | 0.053 | 10.1 | 0.7 |
| **435794** | 2 | 1.69 | 4.1 | 1824 | 9.5 | 0.01 | 4 | 7.7 | 0.8 | 0.019 | 9.8 | 0.3 |
| **435805** | 2.1 | 1.69 | 3.3 | 1576 | 9.8 | 0.011 | 3.8 | 2.1 | 0.7 | 0.066 | 9.9 | 0.2 |
| **435933** | -3.6 | 1.69 | 4.4 | 1724 | 46.8 | 0.01 | 4.2 | 19.2 | 0.7 | 0.008 | 9.3 | 0.4 |
| **435985** | 1 | 1.69 | 4.6 | 1344 | 6.5 | 0.011 | 3.4 | 60.1 | 0.9 | 0.003 | 9.8 | 1.5 |
| **437388** | 2.4 | 1.69 | 4.3 | 1338 | 47.7 | 0.011 | 3.3 | 4.4 | 0.7 | 0.032 | 10.9 | 0.6 |
| **438577** | 2 | 1.69 | 4.6 | 1482 | 3.2 | 0.01 | 3.2 | 2.4 | 0.9 | 0.059 | 8.5 | 0.5 |
| **441582** | 1.1 | 1.69 | 4.6 | 1176 | 34.9 | 0.011 | 4.4 | 27.1 | 0.9 | 0.006 | 7.7 | 4.6 |
| **441736** | 1.3 | 1.69 | 4.3 | 1495 | 9.6 | 0.01 | 3.9 | 44.3 | 0.8 | 0.004 | 8.9 | 6 |
| **441925** | 1.5 | 1.69 | 3.9 | 1718 | 15.9 | 0.011 | 4.5 | 4.1 | 0.8 | 0.035 | 10.2 | 0.6 |
| **442978** | 2.4 | 1.69 | 4.2 | 980 | 19.9 | 0.011 | 4.5 | 2.9 | 0.6 | 0.051 | 10.9 | 0.1 |
| **443108** | 0.6 | 1.69 | 4.5 | 1555 | 3.7 | 0.01 | 3.4 | 3.4 | 0.8 | 0.04 | 9.3 | 0.4 |
| **443176** | 3.2 | 1.69 | 3.8 | 1758 | 54.6 | 0.011 | 4.6 | 4.9 | 0.7 | 0.03 | 10.4 | 0.2 |
| **443240** | NA | NA | NA | NA | NA | NA | NA | NA | NA | NA | NA | NA |
| **444316** | 0.5 | 1.69 | 3.6 | 1706 | 69.9 | 0.011 | 5 | 130.9 | 0.9 | 0.001 | 7.6 | 1.6 |
| **444332** | -0.9 | 1.69 | 2.5 | 1924 | 6.5 | 0.011 | 3 | 1 | 0.6 | 0.124 | 10.4 | 0.1 |
| **444349** | 1.6 | 1.69 | 4.8 | 1665 | 36.7 | 0.01 | 3.8 | 30 | 0.8 | 0.005 | 9 | 0.4 |
| **444391** | 1.3 | 1.69 | 4.3 | 1257 | 0.8 | 0.011 | 3.8 | 8.5 | 0.8 | 0.017 | 9.5 | 0.3 |
| **444446** | 1.9 | 1.69 | 4 | 1720 | 11.4 | 0.01 | 4 | 0.8 | 0.6 | 0.16 | 10.5 | 0.2 |
| **444633** | 0.1 | 1.69 | 4.3 | 1818 | 16.5 | 0.01 | 4.5 | 1.5 | 0.6 | 0.089 | 10.4 | 0.1 |
| **445534** | 2.7 | 1.69 | 4.3 | 1777 | 48.6 | 0.01 | 4.7 | 13.7 | 0.8 | 0.011 | 7.9 | 7.3 |

| | | | | | | | | | | | | |
|---|---|---|---|---|---|---|---|---|---|---|---|---|
| **445535** | 2.7 | 1.69 | 4.9 | 1177 | 1.1 | 0.011 | 3.7 | 0.7 | 0.9 | 0.191 | 9.4 | 3 |
| **445602** | -0.4 | 1.69 | 3.7 | 1725 | 24.4 | 0.011 | 3.3 | 0 | 0.6 | 4.185 | 10 | 0.8 |
| **446645** | 3.5 | 1.69 | 2.7 | 2120 | 22.1 | 0.011 | 4 | 106.2 | 0.7 | 0.002 | 10.2 | 0.6 |
| **449474** | NA | NA | NA | NA | NA | NA | NA | NA | NA | NA | NA | NA |
| **449507** | 0.4 | 1.69 | 4.6 | 1430 | 135.4 | 0.01 | 4.9 | 32.8 | 0.9 | 0.005 | 10.3 | 0.1 |
| **449527** | 3.9 | 1.69 | 3.3 | 2005 | 64.6 | 0.011 | 4.8 | 23.3 | 1 | 0.007 | 9.3 | 0.5 |
| **449614** | 3.6 | 1.69 | 4 | 1895 | 95.6 | 0.011 | 5.1 | 39.1 | 1 | 0.004 | 8.3 | 1.1 |
| **450241** | 2.2 | 1.69 | 4.5 | 1063 | 1.5 | 0.011 | 7.7 | 0.4 | 1.8 | 0.102 | 7.9 | 0.1 |
| **450253** | -0.5 | 1.69 | 5 | 1293 | 89.3 | 0.01 | 7.6 | 0.5 | 1.4 | 0.092 | 8.4 | 3.8 |
| **451146** | NA | NA | NA | NA | NA | NA | 9.2 | 7.8 | 1.6 | 0.009 | 7.6 | 0.7 |
| **451152** | 0.9 | 1.69 | 4.2 | 1382 | 25.6 | 0.011 | 3.3 | 0.3 | 0.8 | 0.421 | 10.6 | 0.8 |
| **451231** | 2.2 | 1.69 | 4.1 | 1736 | 10.6 | 0.011 | 3.5 | 2.5 | 0.7 | 0.053 | 9.6 | 0.4 |
| **451346** | 2.1 | 1.69 | 3.9 | 1689 | 9.4 | 0.011 | 3.7 | 33 | 0.9 | 0.005 | 9.2 | 1.6 |
| **451479** | 4.4 | 1.69 | 4.2 | 1797 | 131 | 0.01 | 5.3 | 94.8 | 0.9 | 0.002 | 9.9 | 0.6 |
| **451709** | -0.4 | 1.69 | 3.1 | 1614 | 57.3 | 0.011 | 7.4 | 0 | 1.4 | 1.359 | 8.6 | 0.5 |
| **453058** | 1.9 | 1.69 | 2.2 | 2252 | 3 | 0.011 | 4.8 | 0.4 | 0.7 | 0.298 | 9.4 | 0.5 |
| **453724** | NA | NA | NA | NA | NA | NA | 0.9 | 180.1 | 0.7 | 0.001 | 8.6 | 6.5 |
| **459589** | 1.7 | 1.69 | 4.3 | 1677 | 42.1 | 0.01 | 8.9 | 0.8 | 1.2 | 0.072 | 7 | 4.4 |
| **459597** | -1.8 | 1.69 | 4.8 | 1379 | 36.4 | 0.01 | 7.7 | 0.5 | 1.5 | 0.109 | 7.9 | 0.3 |
| **461379** | 1.4 | 1.69 | 4 | 1816 | 11.1 | 0.011 | 8.4 | 4.8 | 1.2 | 0.013 | 8.2 | 0.1 |
| **461764** | 1.7 | 1.69 | 3.4 | 1780 | 23.4 | 0.011 | 8.8 | 0.3 | 1.1 | 0.21 | 8.3 | 0.7 |
| **461771** | 1.4 | 1.69 | 4.1 | 1398 | 8.6 | 0.011 | 7.4 | 4.1 | 1.3 | 0.013 | 8.2 | 0.1 |
| **469115** | -0.7 | 1.69 | 3.7 | 1781 | 53.8 | 0.011 | 7.4 | 0.8 | 1.3 | 0.062 | 8 | 0.1 |
| **470855** | -0.5 | 1.69 | 4.5 | 1546 | 100.8 | 0.011 | NA | NA | NA | NA | NA | NA |
| **471467** | -1.7 | 1.69 | 4.4 | 1830 | 70.9 | 0.01 | 8.4 | 0 | 1.1 | 5.634 | 8.1 | 0.1 |
| **471588** | 1.2 | 1.69 | 3.2 | 1882 | 12.9 | 0.011 | 8 | 0.1 | 1 | 0.413 | 8.1 | 0.1 |
| **471890** | 1.8 | 1.69 | 2.3 | 1882 | 7.8 | 0.011 | 8.7 | 4 | 1.4 | 0.016 | 8.3 | 0.2 |
| **473097** | 5.1 | 1.69 | 4.8 | 1793 | 4.4 | 0.01 | 8.7 | 1 | 1.4 | 0.06 | 8.1 | 0.1 |
| **473107** | 2 | 1.69 | 5 | 1441 | 3 | 0.01 | 8.3 | 0.2 | 1.3 | 0.296 | 7.9 | 0.2 |

**Table S6. The fixed parameters and their values in all viral dynamic models.** The initial values of variables are set to 0 except for the two initial values listed below.

| Parameter | Description | Values | Reference |
|---|---|---|---|
| $T_0$ | Total number of (infection free) target cells | $8\times10^7$ cells (nasal) and $1.08\times10^8$ cells (saliva) | See Supplementary Material |
| $E_0$ | Initial number of infected cells | 1 cell | See Supplementary Material |
| c | Virus clearance rate | 10/day | (*2, 3*) |
| k | 1/k is the eclipse period | 4/day | (*2, 3*) |

**Table S7. Model comparison using AIC scores of three models describing the relationship between the viral genome load and cell culture positivity.** In each model, we varied the assumption whether a parameter has random effects or only fixed effects. The lowest AIC score, indicating the best model, is bolded and underlined.

| Model | Parameter assumptions | | AIC |
|---|---|---|---|
| | Random effects | Fixed effect only | |
| Linear model | $A$ | - | 369.6 |
| | - | $A$ | 650.8 |
| Power-Law model | $G, h$ | - | 320.2 |
| | $h$ | $G$ | 318.3 |
| | $G$ | $h$ | 321.7 |
| | - | $h, G$ | 349.2 |
| Saturation model | $J, h, K_m$ | - | 318.1 |
| | $h, K_m$ | $J$ | 318.6 |
| | $J, K_m$ | $h$ | 325.5 |
| | $J, h$ | $K_m$ | <u>**315.6**</u> |
| | $J$ | $h, K_m$ | 322 |
| | $h$ | $J, K_m$ | 330.3 |
| | $K_m$ | $J, h$ | 323.6 |
| | - | $J, h, K_m$ | 345.9 |

**Table S8. The estimated parameters in the best-fit model to the paired RTqPCR and cell culture positivity data, i.e. the saturation model assuming $K_m$ has fixed effect only.** The population estimates of $J, h$ and $K_m$ are 6.6, 0.94 and 4x10$^6$ /mL, respectively.

| ID | Parameter | | | ID | Parameter | |
|---|---|---|---|---|---|---|
| | $J$ | $h$ | | | $J$ | $h$ |
| **432192** | 5.41 | 1.2 | | **444633** | 6.2 | 1.23 |
| **432568** | 2.52 | 0.74 | | **445534** | 9.92 | 0.9 |
| **432662** | 8.86 | 0.96 | | **445535** | 10.56 | 0.82 |
| **432686** | 6.92 | 1.02 | | **445602** | 8.74 | 0.84 |
| **432864** | 7.43 | 1.07 | | **446645** | 5.89 | 1.32 |
| **432870** | 4.24 | 0.43 | | **449507** | 7.03 | 0.55 |
| **433227** | 7.05 | 1.2 | | **449527** | 6.22 | 1.02 |
| **433409** | 2.76 | 0.38 | | **449614** | 5.45 | 1.24 |
| **435729** | 7.48 | 1.06 | | **450241** | 7.93 | 1.35 |
| **435772** | 5.26 | 1.24 | | **450253** | 2.28 | 0.82 |
| **435786** | 2.27 | 0.27 | | **451152** | 6.68 | 1.1 |
| **435794** | 8.67 | 0.74 | | **451231** | 6.95 | 1.02 |
| **435805** | 10.12 | 0.46 | | **451346** | 8.84 | 0.96 |
| **435933** | 7.66 | 0.85 | | **451479** | 6.19 | 0.98 |
| **435985** | 7.4 | 0.48 | | **451709** | 6.86 | 1.36 |
| **437388** | 9.65 | 0.92 | | **453058** | 3.15 | 0.98 |
| **438577** | 6.75 | 1.19 | | **459589** | 5.15 | 1.25 |
| **441582** | 13.72 | 0.76 | | **459597** | 1.45 | 0.76 |
| **441736** | 12.53 | 0.69 | | **461379** | 5.21 | 1.24 |
| **441925** | 5.36 | 1.26 | | **461764** | 2.52 | 1.12 |
| **442978** | 12.31 | 0.62 | | **461771** | 6.14 | 1.53 |
| **443108** | 12.82 | 0.63 | | **469115** | 5.29 | 1.14 |
| **443176** | 5.99 | 1.15 | | **470855** | 4.89 | 1.29 |
| **444316** | 7.44 | 0.67 | | **471467** | 6.02 | 1.25 |
| **444332** | 2.61 | 0.78 | | **471588** | 0.78 | 0.62 |
| **444349** | 10.34 | 0.75 | | **471890** | 2.01 | 0.94 |
| **444391** | 15.29 | 0.7 | | **473097** | 7.59 | 1.2 |
| **444446** | 6.54 | 0.98 | | **473107** | 9.62 | 1.05 |

### Table S9: Viral genotype data

| Participant ID number: | Pangolin_lineage: |
|---|---|
| 432192 | B.1.576 |
| 432568 | B.1.2 |
| 432662 | B.1.2 |
| 432686 | B.1.2 |
| 432864 | B.1.1 |
| 432870 | B.1.2 |
| 433227 | B.1.2 |
| 433409 | B.1.2 |
| 435729 | B.1.582 |
| 435772 | B.1.1.207 |
| 435786 | B.1.2 |
| 435794 | B.1.2 |
| 435805 | B.1.2 |
| 435933 | B.1.2 |
| 435985 | B.1.2 |
| 437388 | B.1.2 |
| 438577 | B.1.311 |
| 441582 | B.1.2 |
| 441736 | B.1.2 |
| 441925 | B.1.2 |
| 442978 | B.1.2 |
| 443108 | B.1.2 |
| 443176 | B.1.2 |
| 443240 | B.1.1.7 |
| 444316 | B.1.577 |
| 444332 | B.1.2 |
| 444349 | B.1.596 |
| 444391 | B.1.2 |
| 444446 | B.1.568 |
| 444633 | B.1.2 |
| 445534 | B.1.2 |
| 445535 | B.1.1.207 |
| 445602 | B.1.2 |
| 446645 | B.1.2 |
| 449474 | B.1.2 |
| 449507 | B.1.2 |
| 449527 | B.1.2 |
| 449614 | B.1.243 |
| 450241 | B.1.1.7 |

| | |
|---|---|
| 450253 | B.1.1.7 |
| 451146 | B.1.1.7 |
| 451152 | C.31 |
| 451231 | B.1.2 |
| 451346 | B.1.1.519 |
| 451479 | B.1.2 |
| 451709 | B.1.1.7 |
| 453058 | B.1.2 |
| 453724 | B.1.2 |
| 459589 | B.1.1.7 |
| 459597 | B.1.1.7 |
| 461379 | B.1.1.7 |
| 461764 | B.1.1.7 |
| 461771 | B.1.1.7 |
| 469115 | B.1.1.7 |
| 470855 | P.1 |
| 471467 | B.1.1.7 |
| 471588 | B.1.1.7 |
| 471890 | B.1.1.7 |
| 473097 | B.1.1.7 |
| 473107 | B.1.1.7 |
